# Wastewater-based estimation of the effective reproductive number of SARS-CoV-2

**DOI:** 10.1101/2021.04.29.21255961

**Authors:** Jana S. Huisman, Jérémie Scire, Lea Caduff, Xavier Fernandez-Cassi, Pravin Ganesanandamoorthy, Anina Kull, Andreas Scheidegger, Elyse Stachler, Alexandria B. Boehm, Bridgette Hughes, Alisha Knudson, Aaron Topol, Krista R. Wigginton, Marlene K. Wolfe, Tamar Kohn, Christoph Ort, Tanja Stadler, Timothy R. Julian

**Author notes:** Corresponding author: Jana S. Huisman; CHN K16, Universitätsstrasse 16, 8092 Zürich, Switzerland. These authors contributed equally (Tamar Kohn, Christoph Ort, Tanja Stadler, Timothy R. Julian). **Competing financial interests** *The authors declare they have no actual or potential competing financial interests*.

## Abstract

**Background:** The effective reproductive number, Re, is a critical indicator to monitor disease dynamics, inform regional and national policies, and estimate the effectiveness of interventions. It describes the average number of new infections caused by a single infectious person through time. To date, Re estimates are based on clinical data such as observed cases, hospitalizations, and/or deaths. These estimates are temporarily biased when clinical testing or reporting strategies change.

**Objectives:** We show that the dynamics of SARS-CoV-2 RNA in wastewater can be used to estimate Re in near real-time, independent of clinical data and without the associated biases.

**Methods:** We collected longitudinal measurements of SARS-CoV-2 RNA in wastewater in Zurich, CH, and San Jose (CA), USA. We combined this data with information on the temporal dynamics of shedding (the shedding load distribution) to estimate a time series proportional to the daily COVID-19 infection incidence. We estimated a wastewater-based Re from this incidence.

**Results:** The method to estimate Re from wastewater works robustly on data from two different countries and two wastewater matrices. The resulting estimates are as similar to the Re estimates from case report data as Re estimates based on observed cases, hospitalizations, and deaths are among each other. We further provide details on the effect of sampling frequency and the shedding load distribution on the ability to infer Re.

**Discussion:** To our knowledge, this is the first time Re has been estimated from wastewater. This method provides a low cost, rapid, and independent way to inform SARS-CoV-2 monitoring during the ongoing pandemic and is applicable to future wastewater-based epidemiology targeting other pathogens.

## Introduction

A critical quantity to monitor an ongoing epidemic is the effective reproductive number Re ^1–4^. Re describes the time-varying average number of new infections caused by a single infectious person throughout the course of their infection. Typically, Re is estimated from case report data (hereafter referred to as R_cc_), including the numbers of new clinical cases, hospitalizations, and deaths ^1, 3–5^. Here, we show that viral RNA concentrations measured in wastewater provide an independent data set to estimate Re (hereafter referred to as R_ww_). This complements existing R_cc_ estimates to provide a more complete picture of transmission dynamics.

Re estimates for SARS-CoV-2 are used to inform regional and national policies ^6, 7^. The Re changes through time and reflects changes in the immune status of the population, policy, climate, and/or individual behaviors ^1, 2^. It can thus be used to estimate the effectiveness of non- pharmaceuticl interventions in disease control ^5, 8–11^. However, R_cc_ estimates have some notable drawbacks. Most importantly, they depend on robust and accurate clinical case surveillance and reporting. Temporal changes in testing capacity, hospitalization criteria, or the definition of COVID-19-related deaths can bias the R_cc_ estimates ^1, 12^. These estimates are also inferred with a delay: R_cc_ is estimable once the infections occurring on that day tested positive and were reported as clinical cases ^1, 2^. This delay differs through time and space, yet is necessary to accurately infer R_cc_, thus complicating the simultaneous computation across geographic regions. Wastewater data may provide an advantage over clinical case data in all these aspects.

SARS-CoV-2 RNA measurements in wastewater can be used to understand COVID-19 epidemiology because infected individuals shed the virus into the sewer system throughout their infection. During the COVID-19 pandemic, SARS-CoV-2 RNA has been repeatedly detected in wastewater and sewage sludge globally ^13–19^, and measured RNA concentrations or loads correlate with clinical case data ^13–15, 17^. Detection of SARS-CoV-2 RNA in the wastewater implies there is at least one actively shedding infected person in the catchment served by the sewer system. Compared to clinical testing, substantially fewer wastewater samples are required to track changes in infection incidence at the community level. Wastewater data has also been integrated into compartmental models of infectious disease transmission, allowing estimation of epidemiological parameters including incidence and the basic reproductive number R0 (which corresponds to the Re in a fully susceptible population at the start of an outbreak) ^20, 21^. These model results are frequently validated against clinical case data, and the good correspondence between both supports the use of SARS-CoV-2 RNA measurements in wastewater to inform disease transmission dynamics. In addition, there are indications that the wastewater may track transmission dynamics more truthfully than cases, especially when test positivity is high ^22^.

Models relating SARS-CoV-2 RNA in wastewater to incidence or transmission rates are driven by assumptions of virus excretion rates into the sewer system. Excretion (via feces, saliva, and/or sputum) varies by individual and through time after infection. Generally, this can be described using a shedding load profile, which captures both the temporal dynamics of shedding (in the shedding load distribution; SLD), and the total amount of virus shed by an infected individual. Clinical studies in various settings have measured shedding from symptom onset onwards. Notable examples include Wölfel et al., who measured virus concentrations in the stool of hospitalized patients ^21, 23^ and Han et al., who included symptomatic and asymptomatic children ^24^. Benefield et al. combined such studies into a systematic review of SARS-CoV-2 viral loads ^25^. However, little is known about shedding prior to symptom onset.

Given uncertainty and variation in estimates of SLDs, modeling approaches to relate wastewater to transmission have varied. For example, Kaplan et al. used an infectivity profile (based on virus concentrations in the upper and lower respiratory tract from Li et al.) rather than information on gastrointestinal shedding to estimate the basic reproductive number R0 from wastewater data ^20, 26^. More work is needed to determine both the SLD and the amount of virus shed during an infection to relate SARS-CoV-2 RNA measurements in wastewater to epidemiology.

We measure SARS-CoV-2 RNA in sewage sludge or wastewater from two distinct monitoring programs (Zurich, Switzerland and San Jose, CA, USA), use the measured RNA to estimate R_ww_, and compare the estimates to R_cc_ obtained from clinical case data. We further determine the SLD that optimizes the fit between R_ww_ and R_cc_ and compare it to previously reported SLDs. We find that R_ww_ is a useful metric to monitor the transmission dynamics of SARS-CoV-2, independent from clinical case data. To our knowledge, this is the first time Re has been estimated from pathogen concentrations in wastewater.

## Methods

### SARS-CoV-2 RNA quantification in wastewater and primary sludge

#### Overall approach

Longitudinal samples of raw wastewater influent from Zurich (Switzerland) and primary sludge from San Jose (California, USA) were collected over several weeks from late 2020 through early 2021. Samples were concentrated, viral RNA was extracted, and SARS-CoV-2 RNA markers as well as pepper mild mottle virus (PMMoV) RNA were quantified in each extract. PMMoV is a plant virus that is found in wastewater at high concentrations and in fairly constant loads, and serves to detect anomalies in the collected sample or problems during concentration and extraction ^27^.

#### Sample collection and processing

##### Zurich approach

From 03 September 2020 to 19 January 2021, 24-hour flow-proportional composite samples of raw influent (after fine screening) were collected from the Werdhölzli wastewater treatment plant (Zurich, Switzerland). Samples were collected twice per week (Thursdays, Sundays) until October 29; afterwards, samples were collected almost daily. Samples were collected in 500 mL polystyrene or polypropylene plastic bottles, shipped on ice, and stored at 4°C for up to 8 days before processing. Samples were processed following the protocol of Fernandez-Cassi et al. 2021 ^22^. Briefly, aliquots (50 mL) were stirred at room temperature for 30 minutes and then clarified by sequential filtration through 2 µm glass fiber pre-filters (Merck) and 0.22 μm SteriCup filters (Merck). The filtrates were concentrated by centrifugation (3000x*g* for 30 minutes) using Centrifugal Filter Units (10kDa Centricon Plus-70, Millipore, USA), followed by concentrate collection from the inverted filter during 3 min at 1000x*g*.

RNA was extracted from concentrates (140-280 μL) using the QiaAmp Viral RNA MiniKit (Qiagen, USA) according to manufacturer’s instructions, using 80 μL of eluate. Until 25 October, samples were processed in duplicate (biological replicates). Samples were extracted once, and a negative extraction control using molecular grade water was run in parallel for every batch of extracted samples.

##### San Jose approach

From 15 November 2020 to 19 March 2021, 125 settled solids samples (approximately 50 mL) were collected and processed daily from the primary settling tank at the San Jose wastewater treatment plant (San Jose, CA, USA) using methods adapted from Graham et al. and described in published protocols ^15, 28–30^. Briefly, 24-hour composite samples were collected in clean plastic containers, immediately stored at 4°C, and transported to the lab for initial processing within 6 hours of collection. The solids were dewatered by centrifugation at 24000*xg* for 30 minutes at 4°C. The supernatant was aspirated and discarded. A 0.5 - 1 g aliquot of the dewatered solids was dried at 110°C for 19-24 hours to determine its dry weight.

Dewatered solids were resuspended in Bovine Coronavirus (BCoV)-spiked DNA/RNA Shield (Zymo Research, Irvine, California, USA), to a concentration of 75 mg/mL. This concentration of solids represented a concentration at which the PCR inhibition of the SARS-CoV-2 assays was minimized based on experiments with solutions containing varying concentrations of solids (see Supplemental Methods)^31^. BCoV was spiked as an external process control. To homogenize samples, 5-10 5/32” Stainless Steel Grinding Balls (OPS Diagnostics) were added to each sample before shaking with a Geno/Grinder 2010 (Spex SamplePrep). Samples were subsequently briefly centrifuged to remove air bubbles introduced during the homogenization process, and then vortexed to re-mix the sample. Samples were either further processed immediately, or stored at 4°C for processing within 7 days.

RNA was extracted from 300 µL of homogenized sample using the Chemagic™ Viral DNA/RNA 300 Kit H96 for the Perkin Elmer Chemagic 360 into 60 µL of eluent followed by PCR Inhibitor Removal with the Zymo OneStep-96 PCR Inhibitor Removal Kit ^29^. Each sample was extracted ten times. In addition, extraction negative and extraction positive controls, consisting of approximately 500 copies of SARS-CoV-2 genomic RNA (ATCC), were extracted using the same protocol as the homogenized samples in each batch of sample extraction.

#### Quantification of viral targets

##### Zurich approach

SARS-CoV-2 N gene markers N1 and N2 were quantified immediately or within one week after RNA extraction (storage at -80°C) using digital RT-PCR (RT-dPCR). RT-dPCR was performed on 5 μL RNA extract as template on either the Bio-Rad QX200 Droplet Digital (01 September 2020 - 7 October 2020) with the One-Step RT-ddPCR Advanced Kit for Probes (Bio-Rad CN 1864021) or Crystal Digital PCR using the Naica System (Stilla Technologies, 8 October - 20 January 2021) with the qScript XLT 1-Step RT-PCR Kit (QuantaBio CN 95132-500). SARS-CoV-2 N1 and N2 markers for the N gene were detected using the 2019-nCoV CDC ddPCR Triplex Probe Assay (Assay ID dEXD28563542, Bio-Rad) according to manufacturer’s instructions, with proprietary primer and probe concentrations. Primer and probe sequences are specified in Table S4, and further dPCR details in Table S5.

For samples processed on the Bio-Rad QX200, 20 µL reaction volumes were prepared in a pre-reaction volume of 22 µL consisting of 5.5 µL of template, 5.5 µL of Supermix, 2.2 µL of Reverse Transcriptase, 1.1 µL of DTT and 1.1 µL of 20x 2019-nCoV CDC ddPCR Triplex Probe Assay. Droplets were generated using the QX100 Droplet Generator (Bio-Rad). PCR was performed on the T100 Thermal Cycler (Bio-Rad) with the following protocol: hold at 25℃ for 3 minutes, reverse transcription at 50℃ for 60 minutes, enzyme activation at 95℃ for 10 minutes, 40 cycles of denaturation at 95℃ for 30 seconds and annealing and extension at 55℃ for 1 minute, enzyme deactivation at 98℃ for 10 minutes, and an indefinite hold at 4℃. Ramp rate was 2℃/second, and the final hold at 4℃ was at least 30 minutes to stabilize droplets. Droplets were analyzed using the QX200 Droplet Reader (Bio-Rad) and thresholding done on the QuantaSoft Analysis Pro Software (Bio-Rad, Version 1.0).

For samples processed on the Crystal Digital PCR, 25 µL reactions were prepared in 27 µL pre-reaction volumes for Sapphire Chips (Stilla Technologies CN C14012) consisting of 5.4 µL of template, 13.5 µL of 2x qScript XLT One-Step RT-PCR, and 1.35 µL of 20x 2019-nCov CDC ddPCR Triplex Probe Assay. Droplet production and PCR were performed on the Naica Geode with the following protocol: reverse transcription at 48℃ for 50 minutes, denaturation at 94℃ for 3 minutes, followed by 40 cycles of denaturation at 94℃ for 30 seconds, annealing and extension at 57℃ for 1 minute. Chips were read and analyzed on the Naica Prism3 using the Crystal Reader and Crystal Miner software (Stilla Technologies).

For the Bio-Rad QX200 samples with more than 12000 droplets with average partitioning volume of 1 nL were deemed acceptable. For the Stilla Crystal Digital PCR 15000 droplets with average 0.8 nL were deemed acceptable. The average (standard deviation) number of droplets observed in samples from QX200 was 15000 (2100) and for the Crystal Digital PCR excluding controls was 24000 (2000). Average copies per partition (relative uncertainty) was 8.7 x 10^-3^ (54%). Technical replicate variability was, on average, less than 20%. The variation amongst distinct RT-dPCR runs (inter-experimental variation) was quantified as the coefficient of variation in the performance of a positive control (100 gene copies (gc)/reaction of synthetic SARS-CoV-2 RNA reference material; EURM-019, Joint Research Center) across 87 runs, and was less than 25%. Assays were only conducted in one laboratory, so reproducibility was not assessed. Example fluorescence plots are provided in the Supporting Information (Figures S11, S12).

Samples were diluted 10-fold in a single step using molecular grade water before quantification in replicate wells. In addition, every thermal cycler run included one positive control and one no template control (NTC) consisting of RNAse/DNAse-free water. Thermal cycle runs and associated samples were deemed acceptable if the NTCs in the run contained 2 or fewer positive droplets, and there was detectable SARS-CoV-2 RNA in the positive controls. All RT-dPCR runs fulfilled these criteria, with an average (standard deviation) concentration of the positive controls of 101 (25) gc/reaction, in line with the target concentration. If the sample concentration was below the limit of quantification (LOQ), an undiluted sample was quantified. The limit of detection (LOD) and limit of quantification (LOQ) of the N1 and N2 markers were determined by processing 10 replicates of synthetic SARS-CoV-2 RNA reference material at target concentrations of 5, 8, 10, 25, 30, and 50 gc/reaction. The LOD was defined as the lowest sample concentration distinguishable from the no template control in at least 8 out of 10 replicates (3 or more positive droplets). At this concentration, there would be a >95% likelihood of detecting the target in at least one of the two technical replicates ^32^. Using this criterion, LOD was determined to be 8 gc/reaction (equivalent to 2560 gc/L wastewater)^32^. LOQ was determined to be 25 gc/reaction (equivalent to 8000 gc/L wastewater), which was the lowest concentration with coefficient of variation less than 25% ^32^. When sample concentrations were below the LOQ, samples were processed without dilution. Only one sample (September 20, replicate B) remained below LOQ in both dilute and undilute samples (22.5 gc/reaction). This sample was included in the analysis anyway.

To test PCR inhibition, the RT-dPCR was repeated using mastermix with a spiked internal positive control consisting of 800 gc/reaction of synthetic SARS-CoV-2 RNA reference material (EURM-019, Joint Research Center) so inhibition testing could be performed on the same assay used for quantification.^33^ Samples were added to the mastermix with a spiked internal control at the same dilution used for quantification of the N1 and N2 markers. If either the observed N1 or N2 concentration in the samples analyzed in mastermix with synthetic SARS-CoV-2 RNA was 80% or less than the sum of the concentration of SARS-CoV–2 RNA in the samples (unspiked) plus the concentration in the sample-free, spiked internal positive control, then the samples were considered inhibited. Inhibited samples were diluted 1:10, retested for SARS-CoV-2 as well as inhibition using the same spiked internal positive control. Dilution sufficiently reduced inhibition for all affected samples.

PMMoV was quantified by RT-qPCR using RNA UltraSense™ One-Step Quantitative RT-PCR System (Applied Biosystems CN 11732927) on a LightCycler® 480 instrument (Roche Life Science, Switzerland) using previously reported primers and probes (Microsynth AG, Switzerland, Table S4)^34, 35^. RNA extract aliquots that were separately stored at -80°C for less than three months were used as template. Samples were prepared in 25 µL reaction volumes consisting of 5 µL of template, 5 µL of 5x Ultrasense Mix, 4 µL of Bovine Serum Albumin (Sigma-Aldrich CN 05470-1G) at 2 mg/mL concentration, 1.25 µL of Reverse Transcriptase, and 1 µL of each primer at final concentrations of 400 nM and 0.25 µL of probe at a final concentration of 250 nM. The RT-qPCR was run with the following program: reverse transcription at 55℃ for 60 minutes, denaturation at 95℃ for 10 minutes, followed by 45 cycles of denaturation at 95℃ for 15 seconds, annealing and extension at 60℃ for 1 minute. PMMoV quantification was performed in six separate RT-qPCR runs by comparison to synthetic DNA standards (gBlock, IDT Technologies) run in duplicate at tenfold dilutions between 5x10^2^ (the lowest concentration measured) and 5x10^7^ per 5 µL reaction. All thermal cycler runs were pooled for analysis. The pooled standard curve had an amplification efficiency of 97.4% and a goodness-of-fit (R^2^) of 0.997.

##### San Jose approach

RNA extracts were used as template in RT-dPCR assays for SARS-CoV-2 N, S, and ORF1a RNA gene targets in a triplex assay, and PMMoV and BCoV in a duplex assay. All primers and probes are listed in Table S4. The SARS-CoV-2 assays were designed using Primer3Plus (https://primer3plus.com/) based on the genome of the severe acute respiratory syndrome coronavirus 2 isolate Wuhan-Hu-1 (Accession Number MN908947.3). The assay was designed to target product size range of 60-200 bp at concentration of dNTPs of 0.8 mM and concentration of divalent cations of 3.8 mM, based on the following optimum (range) conditions: primer size: 20bp (15bp, 36bp); primer melting temperature 60℃ (50℃, 65℃); primer GC content: 50% (40%, 60%); hydrolysis probe size 20bp (15bp, 27bp); hydrolysis probe melting temperature 63℃ (62℃, 70℃); hydrolysis probe GC content: 50% (30%, 80%). The location (length) of the amplicons for N is 28287-28457 (171 bp), S is 23591-23665 (75 bp), and ORF1a is 12885-13063 (179 bp). Cross-reactivity was determined in silico using NCBI Blast. The assays were optimized by varying annealing temperature, and benchmarked against a respiratory virus verification panel using extracted RNA. Limit of the Blank was determined using negative nasal swab samples.

RT-dPCR was performed as previously described for the Bio-Rad QX200 analysis conducted in Zurich using the One-Step RT-ddPCR Advanced Kit for Probes (Bio-Rad 1863021) with primers (900 nM) and probes (250 nM) targeting N, S, and ORF1a RNA. Droplets were generated using the AutoDG Automated Droplet Generator (Bio-Rad). PCR was performed using Mastercycler Pro with the following protocol: reverse transcription at 50℃ for 60 minutes, enzyme activation at 95℃ for 5 minutes, 40 cycles of denaturation at 95℃ for 30 seconds and annealing and extension at either 59℃ (for SARS-CoV-2 assay) or 56℃ (for PMMoV/BCoV duplex assay) for 30 seconds, enzyme deactivation at 98℃ for 10 minutes then an indefinite hold at 4℃. The ramp rate for temperature changes were set to 2℃/second and the final hold at 4℃ was performed for a minimum of 30 minutes to allow the droplets to stabilize.

Droplets were analyzed using the QX200 Droplet Reader (Bio-Rad), with thresholding done using QuantaSoft™ Analysis Pro Software (Bio-Rad, Version 1.0.596). The average (standard deviation) number of droplets in ten merged wells determined from a random subset of ten samples was 176000 (14500). Average (relative uncertainty) of the number of copies per partition in the same subset was 3.2x10^-3^ (52%). As the samples were extracted ten times and each extract analyzed in one well, technical replicate variability incorporates variation from both RNA extraction and RT-dPCR. Sample errors estimated from the merged wells were <10%, in line with coefficient of variation estimates of <8% for all three targets (S, N, ORF1a) in an experiment of replicate (n = 97) positive controls at target concentrations of 400 gc/reaction. Assays were conducted in only one lab, so reproducibility was not assessed. Example fluorescence plots are provided in the associated reference by Topol et al.^30^. All liquid transfers were performed using the Agilent Bravo (Agilent Technologies).

Undiluted extract was used for the SARS-CoV-2 assay template and a 1:100 dilution of the extract (2 µL into 198 µL molecular grade water) was used for the PMMoV and BCoV assay template. The 1:100 dilution is required since PMMoV is in high concentrations, and it is important to be able to quantify the target and not saturate the number of positive partitions.

Each sample was run in 10 replicate wells, extraction negative controls were run in 7 wells, and extraction positive controls in 1 well. In addition, PCR positive controls for SARS-CoV-2 RNA were run in 1 well, and NTC were run in 7 wells. Results from replicate wells were merged for analysis. Negative controls were required to have less than 2 droplets across all wells, PCR positive controls were required to have ∼200 positive droplets, and PCR positive extraction controls were required to have ∼ 50 positive droplets. If controls did not meet these acceptability criteria, then the samples included on that plate were re-processed. Therefore, none of the samples included in this study had controls that failed these acceptability criteria.

#### Data analysis and exclusion criteria

##### Zurich approach

Concentrations of RNA targets were multiplied by the daily flow rate to estimate the total number of genome copies (gc) shed by people within the catchment per day (referred to as loads and reported as gc/day). Samples with PMMoV loads outside the mean plus or minus three times the standard deviation were considered as inconsistent with respect to virus recovery and were excluded from further analysis. Inhibited samples were also removed from further analysis.

##### San Jose approach

Concentrations of RNA targets were converted to concentrations per dry weight of solids in units of gc/g dry weight. PMMoV was also used to monitor virus recovery in the San Jose samples, using the same criteria as for the Zurich samples. BCoV was used to assess virus recovery, and samples were removed from further analysis if the amount recovered was less than 10% of the amount added.

### Deconvolution by the Shedding Load Distribution

To relate the viral RNA loads or concentrations measured in wastewater to the number of new infections per day, we used information on the profile of SARS-CoV-2 RNA shedding into the wastewater by an infected individual in days after infection or symptom onset. In general, this profile contains information about both the magnitude and timing of viral RNA shedding: (i) the SLD ∑*_j_ w_j_* (a unitless distribution which sums to 1) describes the temporal dynamics of shedding, and (ii) a normalization factor *N* describes the total amount of virus shed by an infected individual during the course of infection (in units of gc/infection). After shedding, downstream processes further affect the total amount of viral RNA sampled per infected individual. We assume this does not affect the temporal dynamics, and can be summarised into a second normalization factor M. In general, M will depend on the sewer system, the sampling point within the wastewater treatment plant, choice of sample matrix and processing pipeline. The units of M differ depending on the way viral concentrations were measured: in this study M is unitless for Zurich, and day/g-dry weight for San Jose.

With these definitions, the measurement *C_i_* of viral RNA in the wastewater on day *i* is related to the past incidence of infections *I_j_* on day *j*:

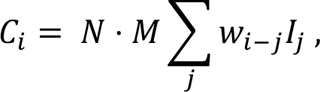

i.e. the observed wastewater measurements are a convolution of the daily infection incidence with the SLD.

To obtain the infection incidence, we first filled gaps in the wastewater data through linear interpolation, and smoothed it using local polynomial regression (LOESS) with 1st order polynomials and tricubic weights that take into account 21 days of data around each point. To deconvolve the resulting time series we used an Expectation-Maximisation algorithm ^2^, which iteratively determines the time series I(t) that maximizes the likelihood of the smoothed wastewater measurements *C̅*(*t*)(either in units of gc/day or gc/g-dry weight), given assumptions on N, M, and ∑*_j_ w_j_*.

For the main analysis, we deconvolved by a SLD which was a combination of the incubation period (the time from infection to symptom onset) and the gastrointestinal SLD from Benefield et al. for the time from symptom onset to shedding ^25^. Figure 3 from Benefield et al. ^25^ was digitized manually, and yielded a gamma distribution with mean 6.7 days and standard deviation 7.0 days ^22^. For the incubation period, we used the distribution of Linton et al.: a gamma distribution with mean 5.3 days and standard deviation 3.2 days ^36^. For additional comparisons (Fig. S5, S6), we exchanged the Benefield distribution for the SLD upon symptom onset reported by Han et al., gamma distributed with mean 4.7 days, standard deviation 1.7 days ^24^, or the symptom onset to death delay distribution from Linton et al., gamma distributed with mean 15 days, standard deviation 6.9 days ^36^.

Since the normalization factors N and M are difficult to measure, and only influence R_ww_ point estimates when off by several orders of magnitude (Fig. S6), we made a simplifying assumption. We assumed the lowest measured RNA load (Zurich) or concentration (San Jose) represents the viral load or concentration from a single infection (N*M). For the Zurich wastewater data, this was 1*10^12^ gc per infection, and for the San Jose sewage sludge measurements this was 2663.7 gc/g-dry weight per infection/day.

### Effective Reproductive Number Estimates

The effective reproductive number was estimated from SARS-CoV-2 RNA loads in wastewater or concentrations in sewage sludge using the pipeline developed in Huisman *et al.* ^2^. In brief, we first transformed SARS-CoV-2 RNA measurements into a time series of infection incidence as described in the Deconvolution section above. Second, we used the R package EpiEstim to estimate the effective reproductive number Re from this infection incidence ^4, 37^. The pipeline further accounts for noise in the observation process, by bootstrapping the observations prior to smoothing and deconvolution. Specifically, we block-bootstrap the log-transformed residuals between the linear interpolated original observations and the smoothed value ^2^.

To estimate R_cc_ for Zurich, we obtained the cases reported for the catchment from the Health Department of Canton Zurich. We then used the pipeline from Huisman *et al.* ^2^, where we deconvolved by a distribution specifying the delay from infection to case confirmation. This was parameterized as the sum of a gamma distributed incubation period with mean 5.3 days, standard deviation 3.2 days ^36^; and a gamma distributed delay from symptom onset to case confirmation with mean 2.8 days, standard deviation 3.0 days (estimated from line list data for canton Zurich, Sep. 2020-Jan. 2021). The reported R_cc_ values for confirmed cases, hospitalizations, and deaths at the cantonal level were taken from https://github.com/covid-19-Re/dailyRe-Data (based on Huisman *et al.*^2^). For the Swiss data, “case confirmation” refers to the earliest recorded date of either a positive test or case reporting.

To estimate R_cc_ in San Jose, we downloaded daily COVID-19 case incidence data for Santa Clara County from the California Health and Human Services Open Data portal (https://data.chhs.ca.gov/dataset/covid-19-time-series-metrics-by-county-and-state). The wastewater from Santa Clara County (population of 1.7 million) is nearly all treated at the San Jose wastewater treatment plant (catchment population of 1.5 million). We estimated R_cc_ using the pipeline from Huisman *et al.* ^2^, with the incubation period as before ^36^; and a gamma distributed symptom onset to case reporting delay distribution with a mean of 4.51 days and standard deviation of 3.16 days (estimated from line list data for Santa Clara County in December; based on personal correspondence with the California Department of Public Health COVID-19 modelling team). During the study period, the mean of this distribution changed from 5.24 to 3.31 days, and the standard deviation from 3.55 to 2.32 days. Negative numbers of cases reported (Dec. 30) were set to 0 for the main analysis (Fig. 2), and to 1000 to test the impact of misreporting (Fig. S2).

To estimate Re for the testing-adjusted cases, we extracted the daily number of positive tests / total number of tests, multiplied by the mean number of tests during the time period (14960.3). This time series was then used to estimate Re, similar to the confirmed cases (with the same delay distribution) ^2^. Technically, the tests are reported by testing date, which typically precedes the reporting date, so this constitutes a misspecification of the delay distribution. However, an analysis where the delay between symptom onset and testing was assumed 0 did not yield qualitatively different results (Fig. S1). We additionally compared our estimates to the R_cc_ estimates for Santa Clara County from the California COVID assessment tool (https://calcat.covid19.ca.gov/cacovidmodels/).

### Comparing Re traces

We assessed how well the Re estimates from SARS-CoV-2 concentrations in wastewater (Rww) match those estimated from case report data (R_cc_) using several measures. First, the average root mean squared error between both point estimates across the time series (“RMSE”):

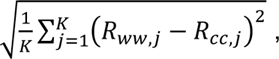

where *j* describes the date, and *K* the length of the time series. Second, the fraction of dates where the R_ww_ point estimate was within the confidence interval of the R_cc_ estimate (“coverage”). Third, the mean average percentage error between the time series (“MAPE”):

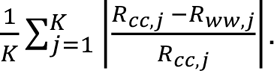

### Scanning across shedding load distributions

To investigate optimal parameters for the SLD, we conducted two separate scans. In the first scan, we varied the parameters of the SLD from infection. In the second scan, we estimated the parameters of the SLD from symptom onset onwards. In the latter case, the delay sampled from the SLD was added to a second sampled delay corresponding to the incubation period (gamma distributed with mean 5.3 days and standard deviation of 3.2 days) ^36^. In both cases, we assumed the SLD was described by a gamma distribution, and varied the mean μ and standard deviation σ on a grid (μ ∈ {0.5, 1.0, …, 15} and σ ∈ {0.5, 1.0, …, 10}). The normalization factor (N*M) was kept fixed to the location-specific value throughout. The R_ww_ for the wastewater data was estimated across 50 bootstrap samples and compared to the R_cc_ for the catchment.

### Availability Statement

All code and case data for Zurich are publicly available through the Github repository https://github.com/JSHuisman/wastewaterRe. Wastewater measurements and daily flow rates for Zurich are available at DOI: 10.25678/0003VC. Measurements from San Jose are available from the Stanford Data Repository https://purl.stanford.edu/bx987vn9177 ^31^, and case data for Santa Clara County is available from the California Health and Human Services Open Data portal (https://data.chhs.ca.gov/dataset/covid-19-time-series-metrics-by-county-and-state).

### Approval

No ethics approval was required for this study as no humans or animals were involved.

## Results

### SARS-CoV-2 RNA in Wastewater

We tracked SARS-CoV-2 RNA concentrations in Zurich, Switzerland and San Jose, California, USA during a rise and fall in clinical COVID-19 cases (Fig. 1A,B; Fig. 2A,B). Data from Zurich were used to develop and assess R_ww_ estimates, and data from San Jose were used to assess the generalizability of the approach.

**Fig. 1:**
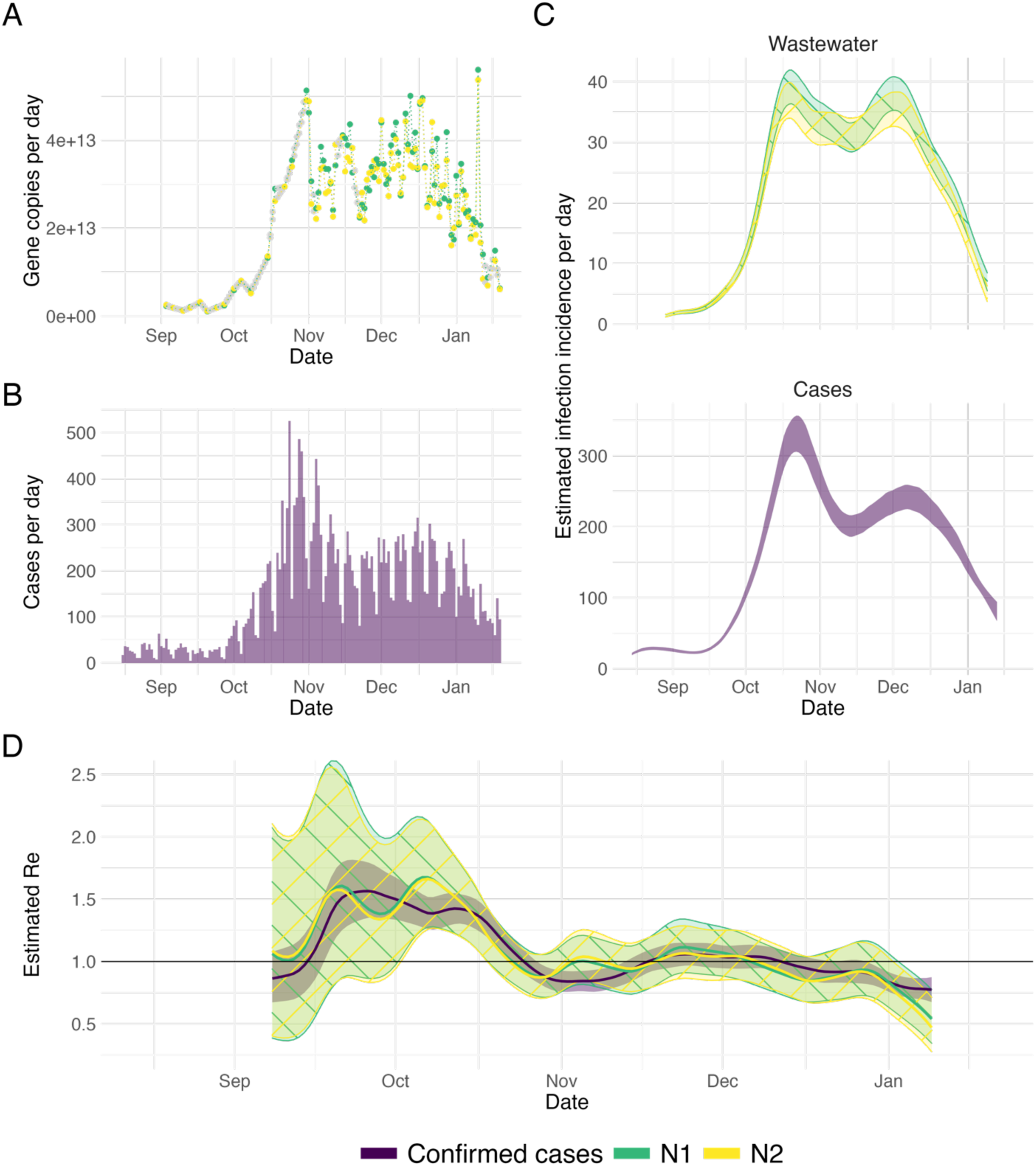
R_ww_ estimation from Zurich (CHE) wastewater measurements. (A) Measured RNA loads of the N1 and N2 markers (green and yellow respectively) between 1 September 2020 and 19 January 2021. Imputed values are indicated in grey. (B) Confirmed cases (purple) in the catchment during the same time period. (C) The estimated infection incidence in the catchment per day from normalized RNA loads of the N1 and N2 markers (top; green and yellow respectively), and case reports (bottom; purple). The measured loads were normalized by the lowest measured value (N*M = 1*10^12^ gc per infection). The ribbons indicate the mean ± standard deviation across 1000 bootstrap replicates. (D) The estimated R_ww_ compared to the R_cc_ from confirmed cases. The colored line indicates the point estimate on the original data, and the ribbons the 95% confidence interval across 1000 bootstrap replicates. The N1 and N2-based confidence intervals nearly completely overlap.

**Fig. 2:**
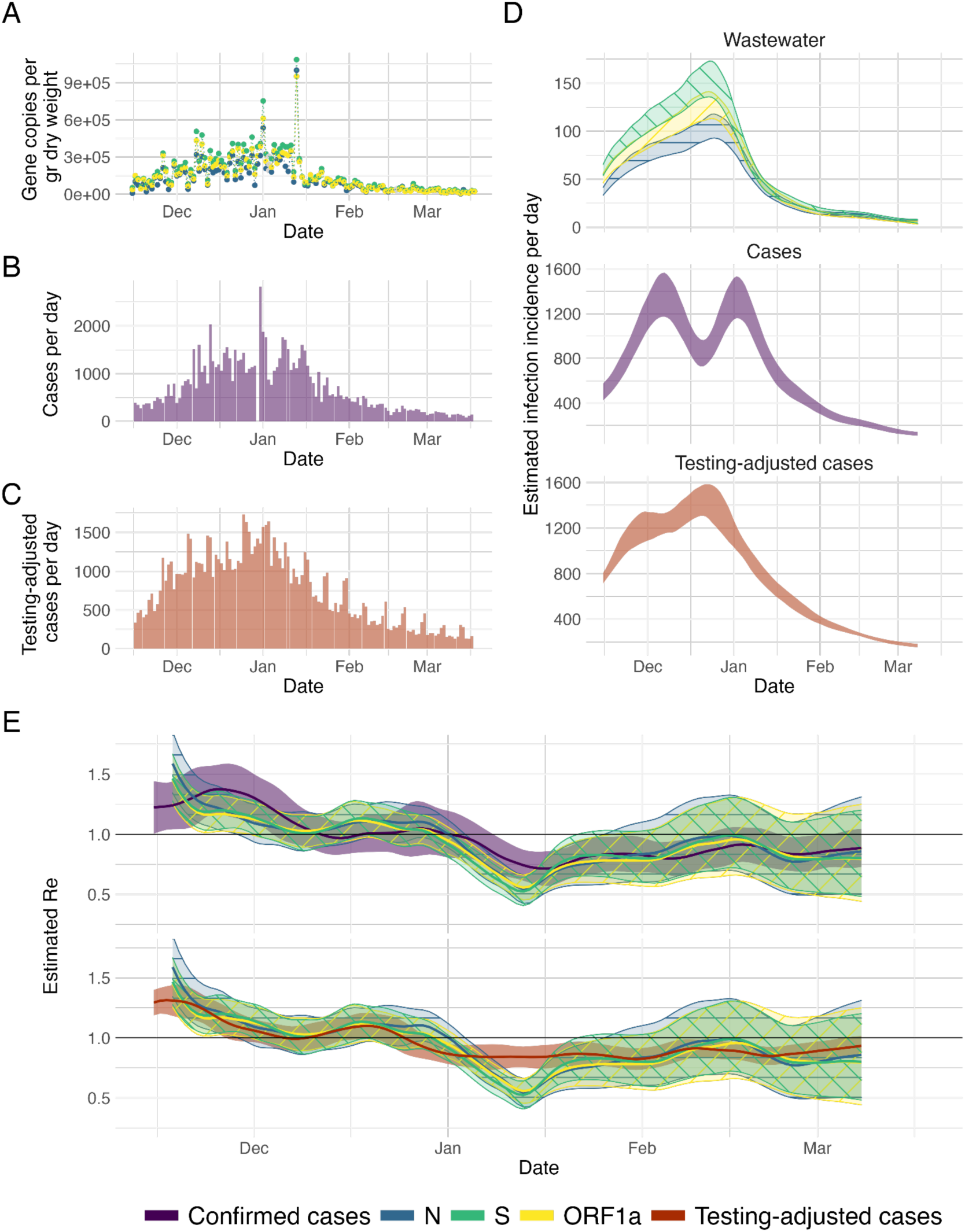
R_ww_ estimation from San Jose (USA) sludge measurements. (A) Measured RNA concentrations of the N, S and ORF1a genes (blue, green, yellow respectively) between 15 November 2020 and 19 March 2021. (B, C) Confirmed cases and testing-adjusted cases in Santa Clara county (purple, red) during the same time period. The testing-adjusted cases describe the number of positive tests / total number of tests per day, normalized by the mean number of tests per day in Santa Clara county during the study period. (D) The estimated infection incidence per day from normalized RNA concentrations (top), case reports (middle) and testing-adjusted cases (bottom). The gene copies per gram dry weight were normalized by the lowest measured value (N*M = 2663.7 gc/g-dry weight per infection/day). The ribbons indicate the mean ± standard deviation across 1000 bootstrap replicates. (E) The estimated R_ww_ compared to R_cc_ from confirmed cases (top) and testing-adjusted cases (bottom). The colored line indicates the point estimate on the original data, and the ribbons the 95% confidence interval across 1000 bootstrap replicates.

**Fig. 3:**
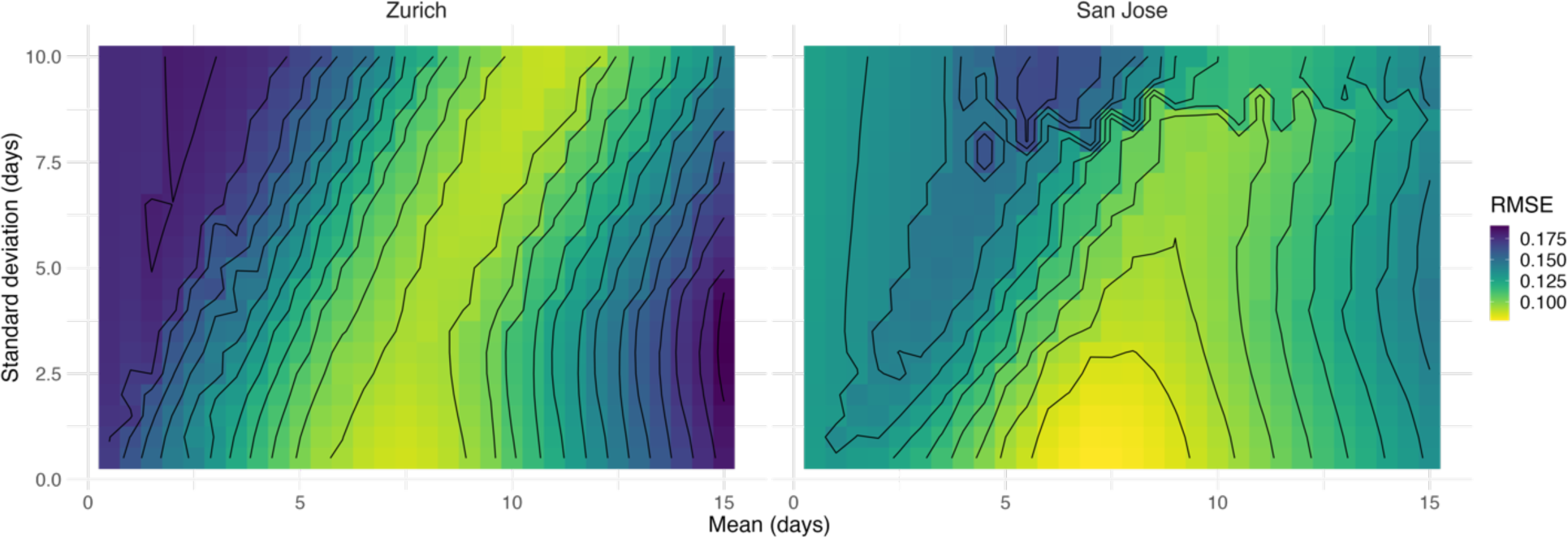
RMSE between R_cc_ and R_ww_ for different shedding load distributions. We scanned across different parameter pairs (mean, standard deviation in days) for the SLD from time since infection. For the city of Zurich, the R_ww_ from N1 loads in wastewater was compared to that of confirmed cases in the catchment. For San Jose, we compared S gene concentrations to confirmed cases in Santa Clara County. The contour lines show SLD parameter pairs with equal RMSE, in steps of 10% of the optimum value.

In Zurich, SARS-CoV-2 N1 and N2 markers of the N gene were detectable in the raw influent samples from the Zurich wastewater treatment plant between 1 September 2020 and 19 January 2021 in all 99 samples collected (Fig. 1A). Of these, the average of the technical replicates was above the limit of quantification in 97, yielding median [range] loads of 13.4 [<12 (limit of detection), 13.7] log10 gc/day (Fig. 1A). Two samples (11 and 29 October 2020) were excluded based on quality control, which included monitoring sample inhibition and consistency of effluent pepper mild mottle virus (PMMoV) loads.

In San Jose, SARS-CoV-2 N, S, and ORF1a genes were quantifiable in the settled solids of the primary settling tank in all 125 samples collected between 15 November 2020 and 19 March 2021 (Fig. 2A). The median [range] concentrations were 4.9 [3.4, 6.0], 5.0 [3.9, 6.0], and 5.0 [3.8, 6.0] log10 gene copies per gram dry weight (gc/g-dry weight) for N, S, and ORF1a genes, respectively (Fig. 2A). Three samples (03 January, 18 February, 19 March 2021) were excluded based on quality control using consistency of PMMoV concentrations.

Details pertaining to PMMoV measurements in all samples and resulting sample exclusion are given in the Supplemental Results.

### Inferring the infection incidence dynamics

Next, we related the RNA measurements in wastewater to the original infection incidence by applying a deconvolution with the shedding load distribution. SARS-CoV-2 wastewater measurements reflect the cumulative contributions of all infected individuals actively shedding virus into the wastewater. The amount of virus shed by each individual varies through time after infection, and is captured in the shedding load profile. In general, this profile contains information about the timing of viral shedding - the shedding load distribution (SLD; which sums to 1) - and the total amount of virus shed - captured by a normalization factor N. To estimate the true number of infections in the sewer shed it is important to estimate the exact value of the normalization factor N, as well as a factor M describing losses along the way from shedding to sample processing. However, to estimate Re it suffices to know the temporal dynamics of shedding and infection (described in more detail in the Methods). As a first approximation, we assumed individuals do not shed prior to symptom onset, and thereafter shed according to the gastrointestinal SLD reported by Benefield et al. ^25^. With this assumption, we found that the dynamics of infection incidence inferred from wastewater measurements in Zurich are similar to the dynamics inferred from clinical case data (Fig. 1C). In particular, both data sources show a steep increase starting from mid-September, and capture two peaks (indicative of Re=1) around late October and early December, each of which is followed by relatively rapid decline in daily case incidence. We later test the sensitivity of our results to the assumed SLD and normalization.

### Estimating the effective reproductive number R_ww_ from wastewater measurements

We used the inferred time series of infection incidence from SARS-CoV-2 RNA measured in wastewater to estimate R_ww_ in Zurich (Fig. 1D). The N1 and N2 markers result in nearly identical R_ww_ estimates, and there is a good correspondence between R_ww_ and R_cc_. Both estimates show a rapid increase up to Re = 1.5 in mid-September, a decline to below 1 in late October, followed by a period where Re was slightly above 1 until dropping more clearly below 1 from early December onwards. R_ww_ and R_cc_ are changing in similar ways, with R_cc_ lagging the R_ww_ trajectory. Since both estimates describe the same underlying epidemic, this suggests that the wastewater measurements may be deconvolved too far back in time (the mean of the SLD is too high), or that the confirmed cases are not deconvolved back far enough (the mean of the delay distribution is too low).

Over the entire time period, the average root mean squared error (RMSE) between R_ww_ and R_cc_ is 0.11 and 0.12 for N1 and N2 respectively. This is smaller than the RMSE between the Re estimates based on different sources of case report data: 0.13 between confirmed cases and hospitalizations; and 0.26 between confirmed cases and deaths (estimated on case report data from canton Zurich, which has a population 3.4x the size of the catchment, for the same time period as R_ww_).

### Application to an independent data source and different wastewater matrix

To test whether these results could be generalized to different geographic locations and wastewater matrices, we analyzed daily-sampled primary sewage sludge data from the San Jose wastewater treatment plant in California.

In San Jose, the inferred infection incidence curves between confirmed cases and wastewater data follow similar trends (Fig. 2D). The inferred incidence from confirmed cases rises rapidly, reaching a maximum and fluctuating at a plateau throughout December. This fluctuation seems primarily caused by reporting errors on Dec. 30th, as it fully disappears when replacing the 0 cases reported that day by 1000 (Fig. S2). Instead, wastewater estimates continue to rise more gradually throughout December, similar to the cases adjusted for test-positivity. Starting in late December, all traces show a similar decrease.

We found that R_ww_ agreed with R_cc_, although there is again some temporal lag between both, which seems more pronounced in November/December than in the second half of the time series (Fig. 2D). The Re estimates based on the testing-adjusted cases are more comparable to R_ww_, both in terms of slope (especially in December) and a more uniform delay throughout the entire time period. This further suggests that the R_cc_ estimates are biased by underreporting and increased testing delays, and not as trustworthy as R_ww_ in this case. A comparison between R_cc_ estimated using different methods (as reported on the website of the California State Department of Public Health) shows substantially larger differences than between the wastewater and the confirmed case estimates from the same pipeline (Fig. S3).

### Minimal Frequency of Wastewater Sampling needed to inform R_ww_

While designing wastewater-based epidemiology studies, an important cost-benefit trade-off centers around the frequency of sampling. We subsampled the daily sampled wastewater measurements in Zurich and San Jose, prior to the Re estimation pipeline, to determine how this would affect the estimated R_ww_. We assessed a range of sampling strategies that differed in the number and identity of the days sampled (e.g. Mon-Wed-Fri or Tue-Fri). For Zurich, we restricted ourselves to the period with daily sampling (22 November 2020 to 11 January 2021). Using the RMSE to quantify the similarity between different R_ww_ estimates, we found that subsampling down to 3 measurements per week still leads to results comparable to a daily sampling regime (Fig. S4, Table S1, Table S2). However, below this frequency the representativity of the R_ww_ estimate starts to depend on which days were sampled.

### Susceptibility of R_ww_ estimates to the shedding load distribution

We showed that Re can be estimated from RNA measurements in wastewater, given an assumption for the SLD. However, there is substantial variation between SLDs described in the literature, across patients, bodily fluids, and geographic locations. The shape of the SLD, in particular the mean of the gamma distribution used, affects the inferred timing of peak infection incidence (with larger means shifting the incidence further back in time; Fig. S5). In our pipeline, we also observe that smaller normalization factors N*M increase the amplitude of the estimated R_ww_, albeit only when mis-specified by more than 5 orders of magnitude (Fig. S6). In principle, the inference of the Re point estimate from an infection incidence is independent of the magnitude of this incidence ^2, 4^. However, the expectation maximization algorithm used for deconvolution in our pipeline was optimized for data on the scale of infections per day. Here, we have chosen to normalize the wastewater measurements such that the considered gene loads are on that same scale, since R_ww_ otherwise reacts too strongly to changes in the daily incidence.

### Estimating the shedding load distribution from the fit between clinical and wastewater data

Instead of assuming a single SLD and estimating R_ww_ based only on that distribution, we also asked which SLD would maximize the similarity between the R_ww_ and R_cc_ estimates. We numerically scanned across different SLDs and quantified the resulting goodness of fit between the R_ww_ and R_cc_ for both Zurich and San Jose. We assumed the SLD is described by a single gamma distribution, starting at infection, and searched for the optimal fit on a grid of mean-standard deviation parameter pairs (Table 1, Fig. 3). The fit was quantified using the root mean squared error (RMSE), coverage, and mean absolute percentage error (MAPE). Since the measurements of the different genetic markers followed nearly identical patterns in both locations (Figs. 1 and 2), we conducted the SLD optimization analysis only for the N1 marker in Zurich, and S gene in San Jose.

**Table 1:** Parameters of the optimal shedding distribution from infection. We scanned across different (mean, standard deviation) parameter pairs for the SLD from time since infection. For Zurich, the Re from N1 loads in wastewater was compared to that of confirmed cases in the catchment. For San Jose, we compared S gene concentrations to confirmed cases in Santa Clara County. For all values of the scan see Figs. 3, S7, S8. All parameters are in units of days. For the coverage, the 95% confidence intervals of R_cc_ and R_ww_ were based on 50 bootstrap replicates in each comparison.

| Comparison Method | Optimal pair (mean; sd) | Mean within 10% from the optimum | SD within 10% from the optimum |
| --- | --- | --- | --- |
| RMSE (Zurich) | (7.5; 0.5) | [6, 11.5] | [0.5, 10] |
| Coverage (Zurich) | (7.5; 0.5) | [6, 12.5] | [0.5, 10] |
| MAPE (Zurich) | (11; 9.5) | [6.5, 11] | [0.5, 10] |
| RMSE (San Jose) | (7.0; 0.5) | [6, 9] | [0.5, 3] |
| Coverage (San Jose) | (5; 0.5) | [1, 11] | [0.5, 10] |
| MAPE (San Jose) | (6; 0.5) | [5, 8] | [0.5, 2.5] |

The optimal fits based on these metrics suggest that the SLD has a mean between 7-11 days in Zurich and 5-7 days in San Jose, with a very low standard deviation of 0.5 days in both locations (Table 1). However, there is some non-identifiability in our analysis, with most optimal value pairs lying along a ridge (Fig. 3; repeated for coverage: S7, and MAPE: S8). This ridge corresponds to SLDs with a similar median, which result in nearly indistinguishable R_ww_ estimates (examples shown in Fig. S9). If we consider the parameters yielding a fit within 10% from the optimum, the parameter ranges found in both locations are compatible and jointly suggest an SLD with mean between 6-9 days, and standard deviation between 0.5-3 days. Longer time series and more locations would further constrain this distribution. Compared to the delay between infection and case reporting, the SLD introduces a similar or lower mean delay to R_ww_. For Zurich, the cases were delayed with respect to infection by 8.1 days on average, which is comparable to the 6-9 days for R_ww_. For San Jose, instead, the delay distribution of the case report data had a mean of 9.8 days. There, the wastewater may lead the confirmed cases by 1-4 days, if the current testing and reporting regime is upheld.

To compare against published SLDs, which are frequently parameterized from symptom onset instead of infection, we conducted a second analysis. Here we assumed individuals do not shed during their incubation period and subsequently shed with a gamma distribution, starting at symptom onset. In this case, we find optimal SLDs with a mean between 0.5-3 days for San Jose and 3.5-5.5 days for Zurich (Table S3, Fig. S10). These optimal distributions have a lower mean than the SLD reported by Benefield et al. (mean 6.7 days), and Han et al. (mean 4.7 days) ^24, 25^. If we add the mean incubation period (5.3 days) to the results of this scan, we find that for both locations the mean delay between infection and shedding is comparable to the mean of the SLD we estimated from infection.

## Discussion

We showed that regular measurements of SARS-CoV-2 concentrations in wastewater and settled solids can be used to estimate the effective reproductive number Re. The difference between Re estimates from wastewater (Rww) and from case report data (R_cc_) is similar to the difference between Re estimates based on different types of case report data (clinical cases, hospitalizations, and deaths). This did not depend on which of the measured gene targets was used to estimate R_ww_. We further showed wastewater samples should be collected at least 3 times per week to reliably estimate past R_ww_, in line with analyses based on direct comparison of wastewater signals to clinical cases ^15, 38^. For real-time monitoring of R_ww_, more frequent measurements may be preferable to ensure stable estimates when new data comes in.

Estimating R_ww_ requires accurate characterization of the SLD, i.e. the temporal dynamics of shedding. In our primary analysis, we used the distribution for gastrointestinal shedding from Benefield et al. ^25^. In using this SLD, we implicitly assumed that fecal shedding dominates the viral load in wastewater. However, there is a wide range in published viral shedding loads, and it is unclear which - if any - accurately capture viral shedding dynamics of people within a catchment. Virus shed in saliva, sputum, and feces are possible contributors to the total amount of virus RNA in the wastewater ^39^. While upper respiratory tract swabs show peak viral loads around the day of symptom onset, there are indications that sputum samples peak a few days later, and feces even after that ^40–42^. Studies differ in the inferred timing of peak viral load (even in the same bodily fluids), and there is a general lack of information to constrain dynamics prior to symptom onset ^43^. Additionally, the duration and magnitude of viral shedding seem to differ within different populations (for example, due to age or severity of disease) ^44, 45^. However, these individual differences will probably average out in a sufficiently large catchment and better estimates of the SLD are likely to become available as prospective sampling studies report results.

We showed that the optimal SLD can also be inferred from the fit between R_ww_ and R_cc_. Once the SLD has been estimated from historic wastewater and case data, it may from then on provide a more accurate estimation of R_ww_, than using one of the published SLDs. Indeed, here we show a range of gamma-distributed SLDs inferred from our wastewater data that generally align with, but have lower means than published SLDs based on patient shedding profiles. Optimization based on alignment between R_ww_ and R_cc_ assumes accuracy of R_cc_, which only holds when there is adequate clinical case surveillance. However, given widespread wastewater monitoring coincident to clinical case reporting, broader application of our methods would help constrain the SLD of SARS-CoV-2.

The utility of wastewater measurements for Re estimation is independent of the pipeline used to estimate Re. Here, we report results obtained with the pipeline of Huisman et al. ^2^. However, many estimation methods exist, differing in assumptions on smoothing, deconvolution, uncertainty quantification as well as the underlying method to estimate Re from infection incidence ^1, 3, 7, 46, 47^. Although the Re point estimate is not affected by the absolute magnitude of the infection incidence (and thus comparable across wastewater treatment plants with differing sampling protocols), the rest of our pipeline (in particular the deconvolution) was originally developed specifically for use with clinical data. Thus, we had to normalize the measured wastewater concentrations to the same order of magnitude as the case data. Further development could make the method more specifically adapted to wastewater data, and alleviate this dependence on the normalization.

Estimates of R_ww_ are independent of biases influencing clinical case-based estimates. R_cc_ estimates are based on only the subset of infections, hospitalizations, and/or deaths that are captured by surveillance within the healthcare system. If this subset changes, for instance due to developments in testing or reporting policy, the resulting R_cc_ estimates will be temporarily biased ^2, 4^. In Geneva, Switzerland, seroprevalence studies showed that the number of infections per reported case varied substantially, from an estimated 11.6 infections per reported case as of May 2020 to only 2.7 as of December 2020 ^48, 49^. During that period, SARS-CoV-2 RNA concentrations in wastewater better reflected the dynamics than the clinical cases ^22^.

However, R_ww_ estimates are also prone to biases. People’s behaviors, such as defecation timing outside of a daily routine ^50^ and/or movement into or out of the catchment ^51^ can influence R_ww_ estimates, particularly when the number of infected individuals is low. RNA signals may also be impacted during sewer transport, with persistence influenced by environmental conditions (i.e., temperature) and/or sewage composition (i.e., solids content) ^52–55^. Furthermore, sample processing required to quantify SARS-CoV-2 RNA may introduce variation, as suggested by substantial day-to-day variation in measurements ^13, 15, 22, 56^. Finally, R_ww_ estimates are informed by the number and proportion of infected and/or shedding people within the catchment: if there are too few active shedders, R_ww_ may be very sensitive to the increased fluctuations in SARS-CoV-2 RNA concentrations.

To conclude, deriving Re from wastewater offers an independent method to track disease dynamics. Wastewater-based epidemiology is used globally to track the COVID-19 pandemic (https://www.covid19wbec.org/covidpoops19) ^13–19^. The data collected within these campaigns could be used to estimate R_ww_ with a robust method, not influenced by heterogeneous testing and reporting strategies, and hence more comparable across geographic areas. Additionally, R_ww_ estimates could be derived for the transmission of SARS-CoV-2 variants and/or other pathogens for which SLDs are known. SARS-CoV-2 variants, including Variants of Concern (VOCs), are readily detectable in wastewater ^57–59^ as are other pathogens (e.g., norovirus, enterovirus, hepatitis A) ^60–62^. This could provide the temporal, quantitative wastewater measurements needed to estimate R_ww_. Wastewater surveillance allows estimating R_ww_ to track disease transmission dynamics in near-real time, using low cost, rapid, and geographically-comparable methods, and can be used when reporting clinical cases is not feasible, mandatory, or much delayed compared to infection and shedding.

## Data Availability

All code and case data for Zurich are publicly available through the Github repository https://github.com/JSHuisman/wastewaterRe. Wastewater measurements and daily flow rates for Zurich are available at DOI: 10.25678/0003VC. Measurements from San Jose are available from the Stanford Data Repository https://purl.stanford.edu/bx987vn9177, and case data for Santa Clara County is available from the California Health and Human Services Open Data portal (https://data.chhs.ca.gov/dataset/covid-19-time-series-metrics-by-county-and-state).

https://doi.org/10.25678/0003VC

https://purl.stanford.edu/bx987vn9177

## Acknowledgements

We thank members from the Bonhoeffer and Stadler groups for helpful discussions, Carola Bänziger (Eawag), Claudia Scheckel (Oncobit AG, Switzerland), and Bruno Mueller and Sergey Yakushev (Microsynth AG, Switzerland) for assistance with and/or knowledge exchange on method development. We further thank the operators of the Zurich WWTP for providing samples; Alexander J. Devaux and Charlie Gan for their help in analyzing the Zurich samples; the staff of the San Jose WWTP including Payak Sarkar, Noel Enoki, and Amy Wong; the Health Department of Canton Zurich for catchment-specific case numbers; and the California Department of Public Health Covid-19 modelling team for input on the Re estimates and symptom onset to case confirmation delay distribution for the state of California. We thank the reviewers for their suggestions to improve the manuscript. XFC, TS, CO, TRJ and TK acknowledge funding from the Swiss National Science foundation (Special Call on Coronaviruses; 31CA30_196267 and 31CA30_196538). CO, TRJ and TK further acknowledge discretionary funding from Eawag and EPFL. XFC was a fellow of the European Union’s Horizon 2020 research and innovation program under the Marie Skłodowska–Curie Grant Agreement No. 754462. The San Jose wastewater data acquisition and curation was funded by the CDC-Foundation.

## Author contributions

JSH, TK, CO, TS, TRJ conceived the study; JSH developed the analytical framework, designed and performed computational analyses; JS, AS, TS contributed to the analytical methods; LC, XFC, PG, AKu, ES, ABB, BH, AKn, AT, KRW, MKW, TK, CO, TRJ developed experimental protocols and performed wastewater sampling; XFC, ABB, KRW, TK, CO, TS, TRJ supervised the study and secured funding; JSH, TK, TRJ wrote the original draft; all authors reviewed and approved the final manuscript.

## Supplemental Material

**Fig. S1:**
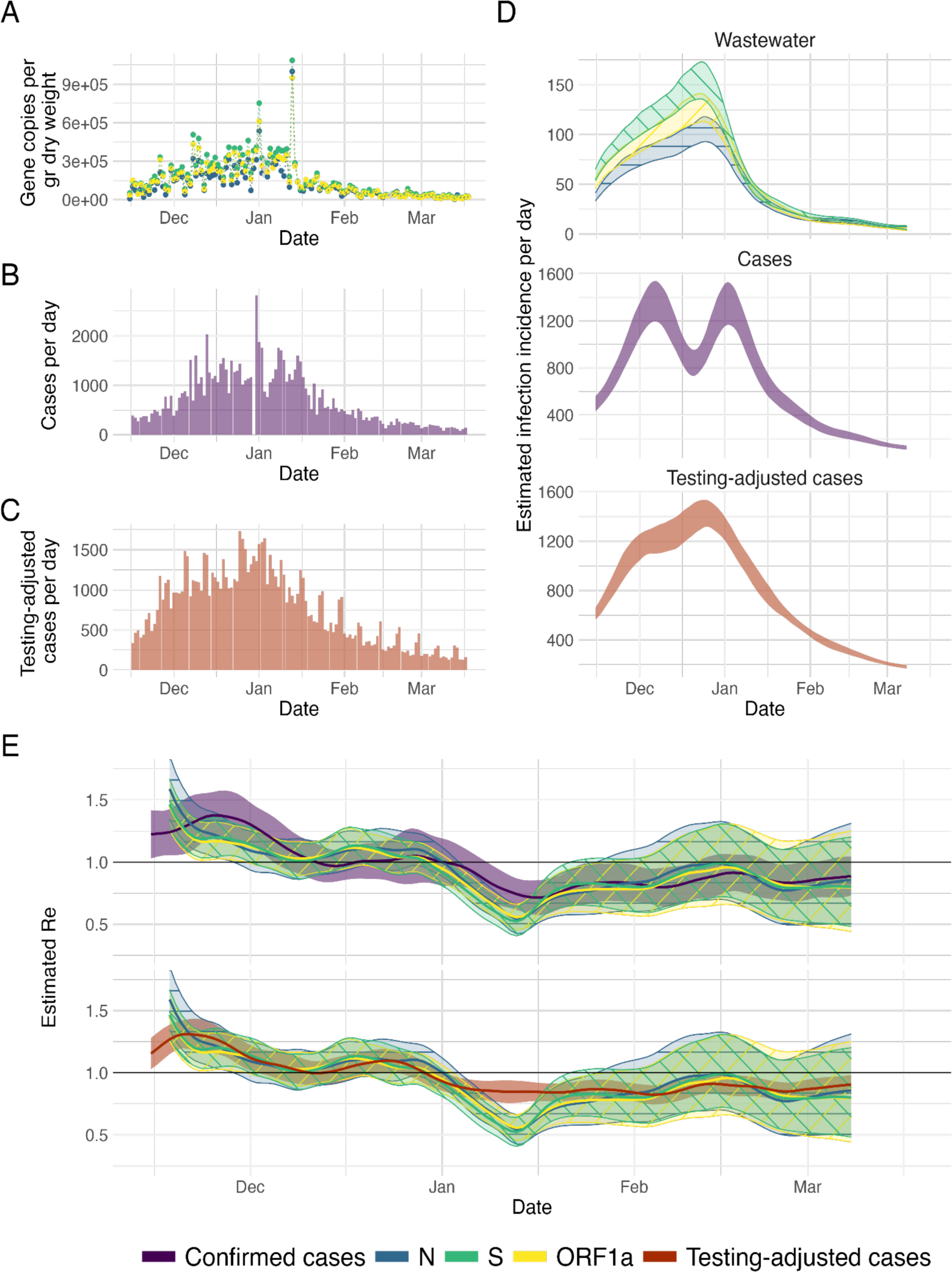
R_ww_ estimation from San Jose (USA) sludge measurements, assuming zero delay between symptom onset and testing: (A) Measured RNA concentrations of the N, S and ORF1a genes (blue, green, yellow respectively) between 15 November 2020 and 19 March 2021. (B, C) Confirmed cases and testing-adjusted cases in Santa Clara county (purple, red) during the same time period. The testing-adjusted cases describe the number of positive tests / total number of tests per day, normalized by the mean number of tests per day in Santa Clara county during the study period. In contrast to Fig. 2 in the main text, the delay between symptom onset and testing was assumed 0, rather than the same as for case reporting. (D) The estimated infection incidence per day from normalized RNA concentrations (top), case reports (middle) and testing-adjusted cases (bottom). The gene copies per gram dry weight were normalized by the lowest measured value (N*M = 2663.7 gc/g-dry weight per infection/day). The ribbons indicate the mean ± standard deviation across 1000 bootstrap replicates. (E) The estimated R_ww_ compared to R_cc_ from confirmed cases (top) and testing-adjusted cases (bottom). The colored line indicates the point estimate on the original data, and the ribbons the 95% confidence interval across 1000 bootstrap replicates.

**Fig. S2:**
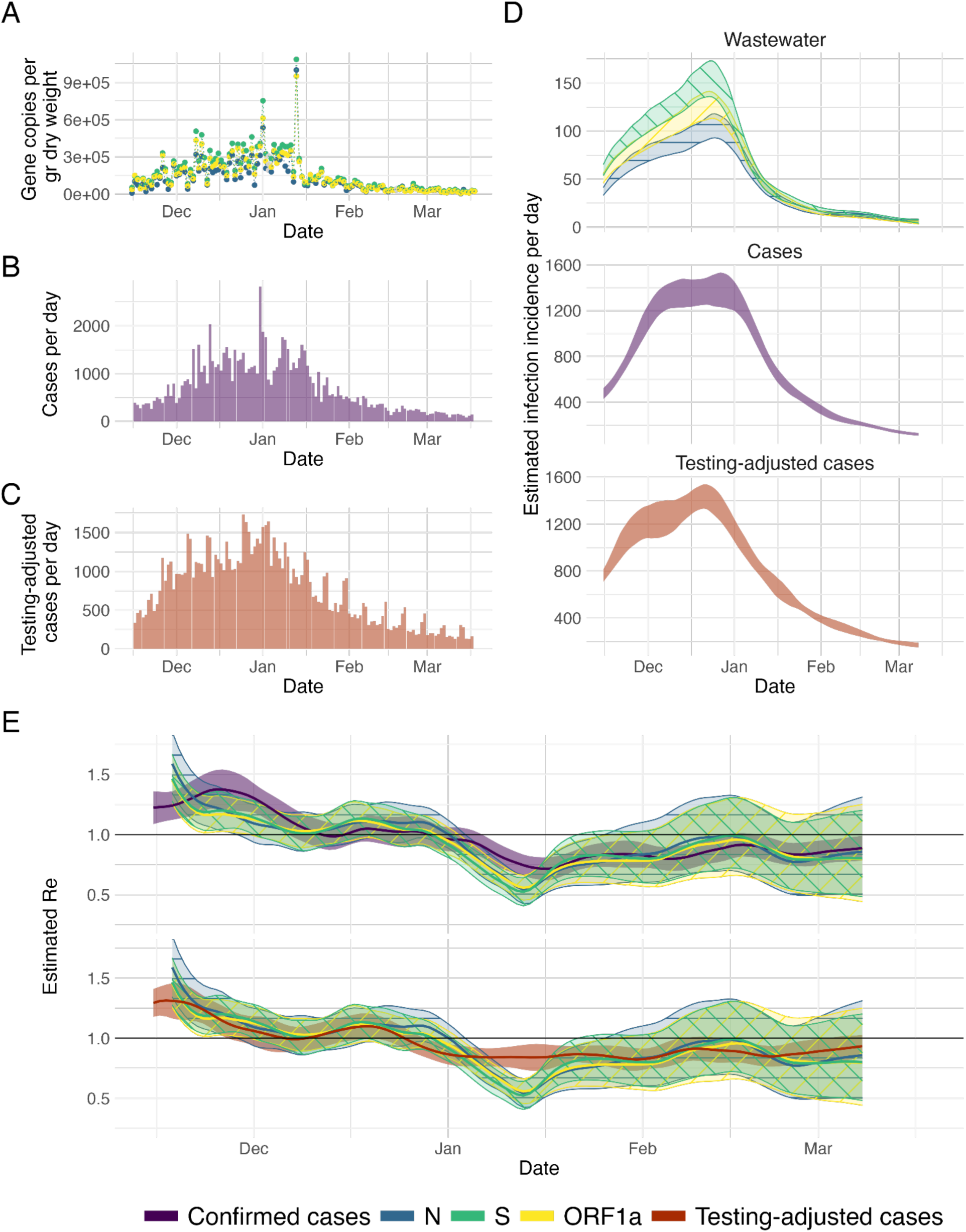
R_ww_ estimation from San Jose (USA) sludge measurements, accounting for case reporting errors: (A) Measured RNA concentrations of the N, S and ORF1a genes (blue, green, yellow respectively) between 15 November 2020 and 19 March 2021. (B, C) Confirmed cases and testing-adjusted cases in Santa Clara county (purple, red) during the same time period. In contrast to Fig. 2 in the main text, the missing cases on Dec. 30th were set to 1000 in panel (B). The testing-adjusted cases describe the number of positive tests / total number of tests per day, normalized by the mean number of tests per day in Santa Clara county during the study period. (D) The estimated infection incidence per day from normalized RNA concentrations (top), case reports (middle) and testing-adjusted cases (bottom). The gene copies per gram dry weight were normalized by the lowest measured value (N*M = 2663.7 gc/g-dry weight per infection/day). The ribbons indicate the mean ± standard deviation across 1000 bootstrap replicates. (E) The estimated R_ww_ compared to R_cc_ from confirmed cases (top) and testing-adjusted cases (bottom). The colored line indicates the point estimate on the original data, and the ribbons the 95% confidence interval across 1000 bootstrap replicates.

**Fig. S3:**
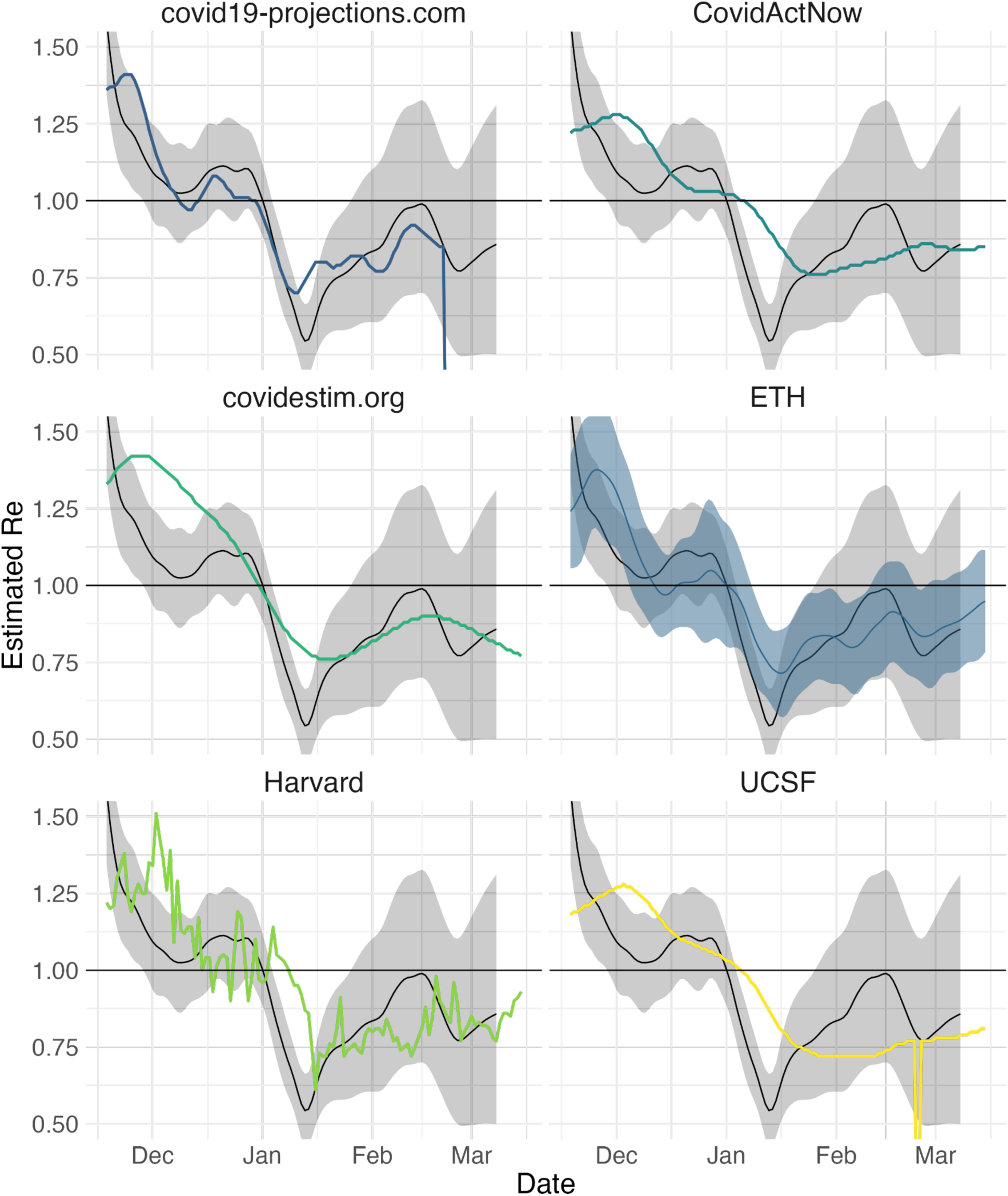
The estimated R_ww_ based on N gene measurements in San Jose (black line, grey 95% confidence intervals) compared to different R_cc_ estimates for Santa Clara county from the California COVID assessment tool (CalCat). The R_ww_ estimate from Fig. 2D was compared to 6 methods to estimate R_cc_, indicated by the color and panel title. “ETH“ refers to the pipeline of Huisman et al. ^2^ applied to the daily confirmed case data from Santa Clara county, i.e. the same comparison as shown in Fig. 2D. We excluded the SEIR model based estimates included on the CalCat website (since these performed visibly worse than the other methods).

**Fig. S4:**
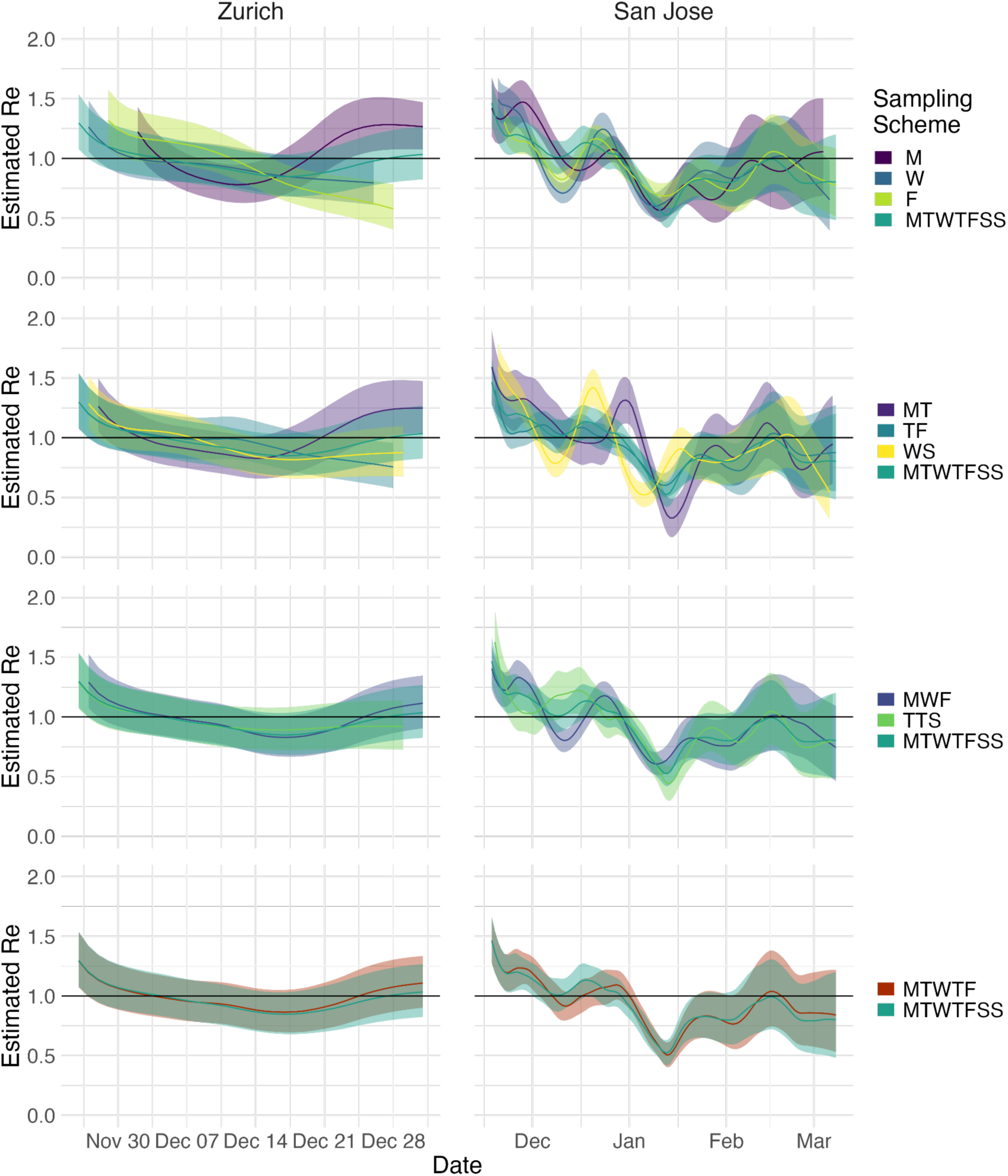
The effect of sampling frequency on the R_ww_ estimates in Zurich and San Jose. The data stems from measurements of the N1 marker for Zurich between 2020-11-22 and 2021-01-11, and the S gene for San Jose between 2020-11-15 and 2021-03-19. The row indicates the number of samples taken per week (1, 2, 3, 5), while the colors indicate the sampling schedule: MTWTFSS corresponds to daily sampling, MTWTF sampling during the working week, MWF Mon-Wed-Fri, TTS Tue-Thu-Sat, MT Mon-Thu, TF Tue-Fri, WS Wed-Sat, M Monday, W Wednesday, F Friday. The colored line indicates the point estimate on the original data, and the ribbons the 95% confidence interval across 1000 bootstrap replicates.

**Fig. S5:**
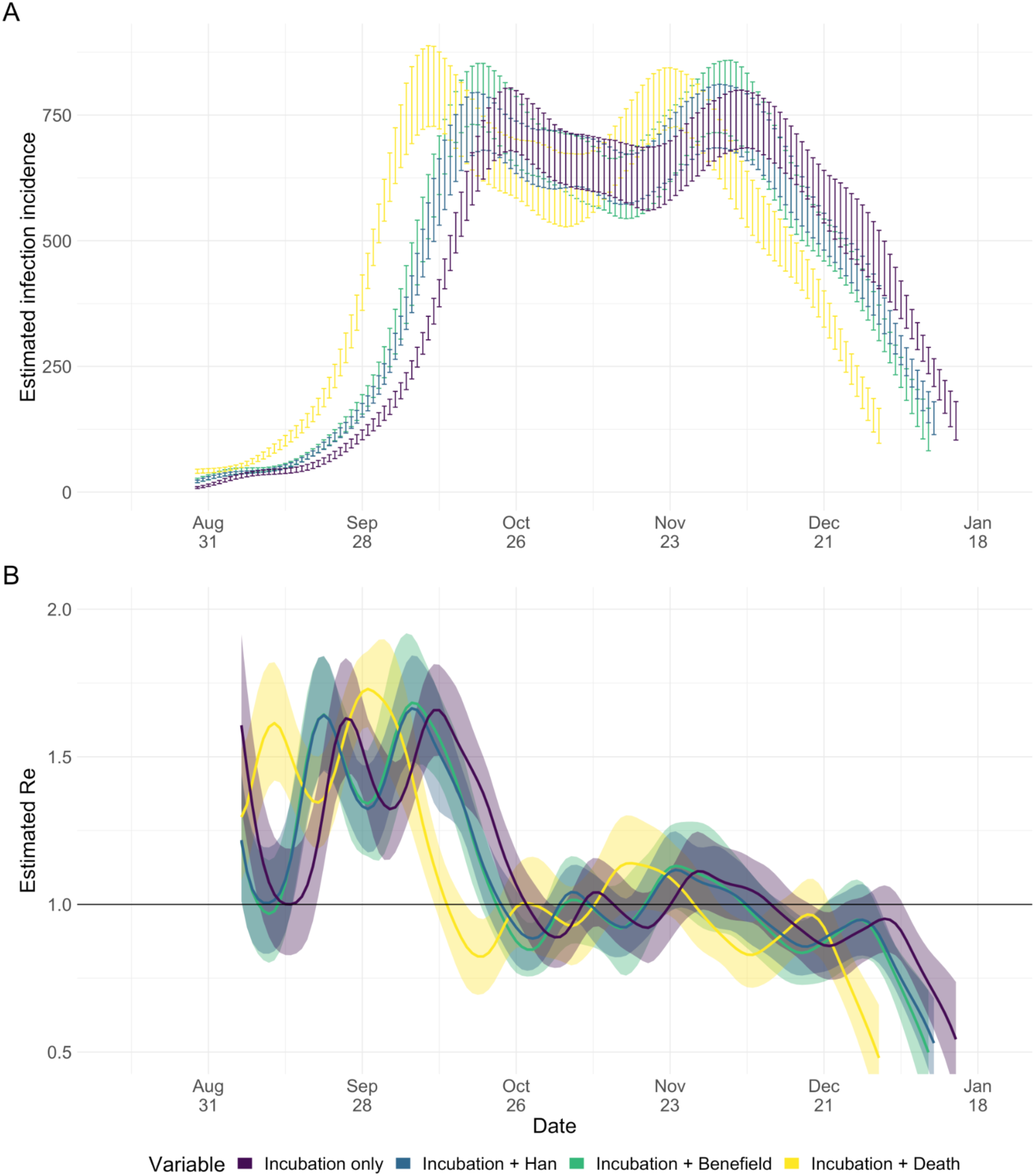
The impact of the shedding load distribution on (A) the inferred infection incidence and (B) R_ww_ estimation from the Zurich wastewater data. The results are compared for four SLDs (see Methods): the Benefield SLD upon symptom onset (Incubation + Benefield) ^25^, the Han SLD upon symptom onset (Incubation + Han) ^24, 25^, shedding only on the day of symptom onset (Incubation only), shedding only on the day of death (Incubation + Death)^36^. The error bars (A; mean ± standard deviation) and 95% confidence intervals (B) are basedon 50 bootstrap replicates per condition.

**Fig. S6:**
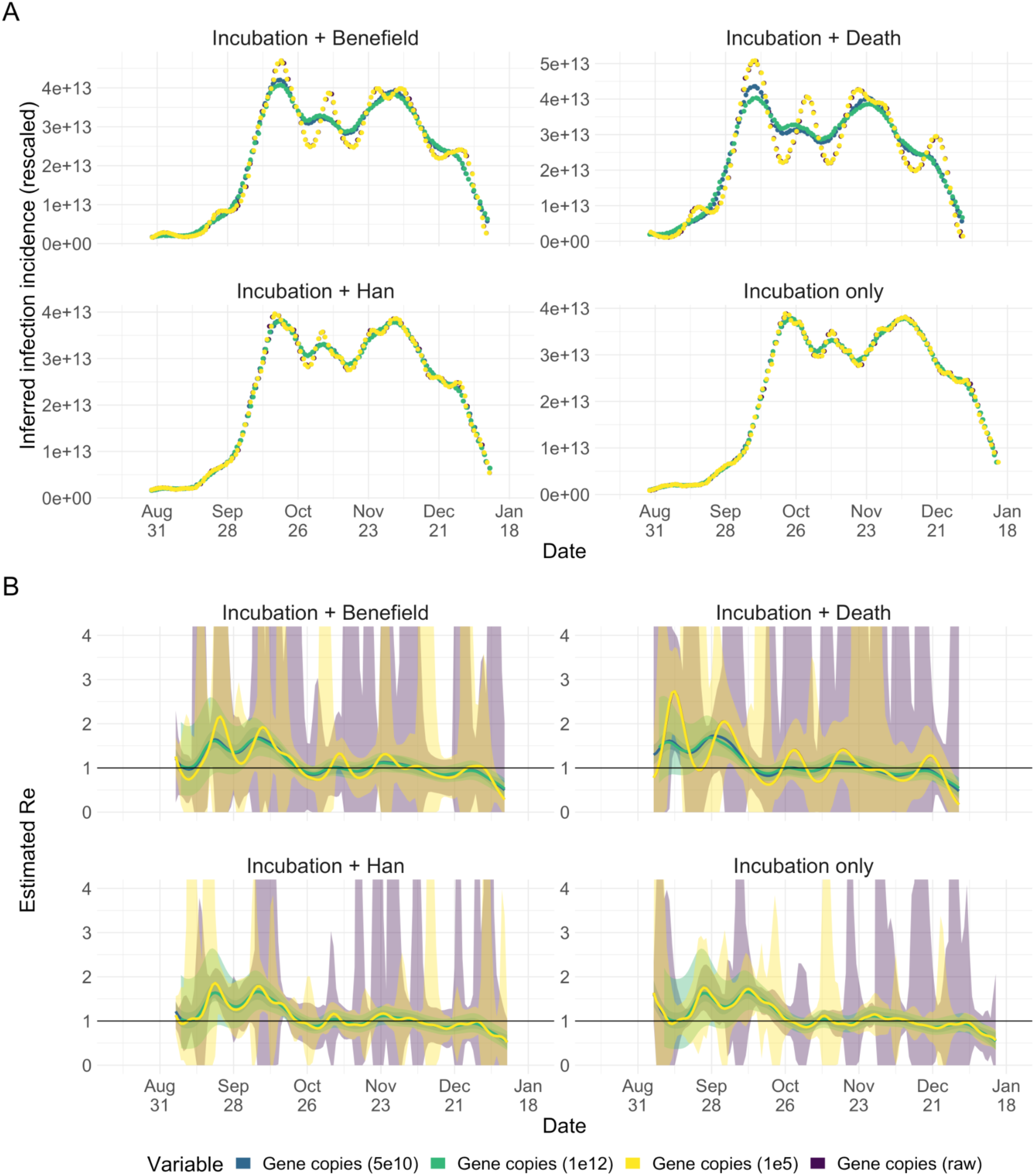
The effect of data normalization on the estimated R_ww_ in Zurich. We calculated R_ww_ from the measured N1 marker indicating the N gene copy loads, normalized in four different ways: by 1x10^12^ gc per infection (the order of magnitude of the lowest measured concentration), by 5x10^10^ gc per infection (roughly rescaled to case incidence), by 1x10^5^ gc per infection (to cover the space of possible orders of magnitude), no normalization. **(A)** After normalization the measurements were deconvolved, and rescaled back to the original magnitude (multiplied by the normalization factor) to illustrate differences in the inferred infection incidence. **(B)** The resulting R_ww_ estimates differ both in the mean (due to the deconvolution illustrated in A) and width of the uncertainty interval. The results are compared for four SLDs (panels; see Methods): the Benefield SLD upon symptom onset (Incubation + Benefield) ^25^, the Han SLD upon symptom onset (Incubation + Han) ^24, 25^, shedding only on the day of symptom onset (Incubation only), shedding only on the day of death (Incubation + Death) ^36^. The 95% confidence intervals (B) are based on 50 bootstrap replicates per condition.

**Fig. S7:**
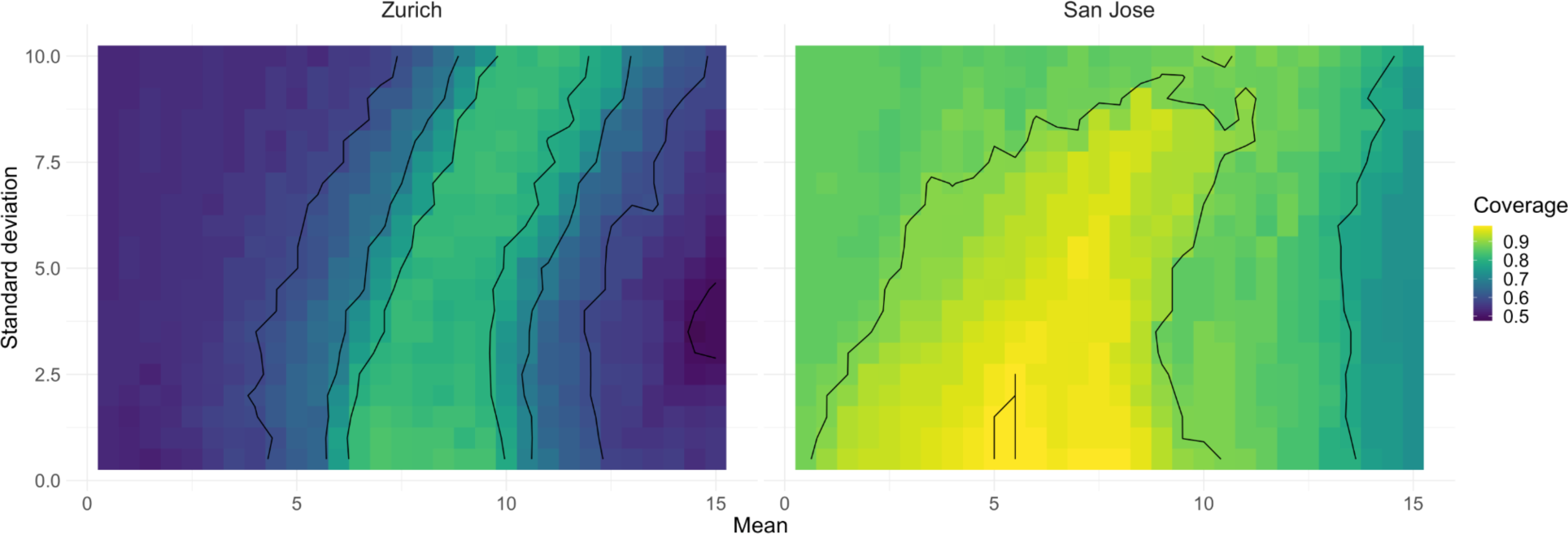
Coverage between R_cc_ and R_ww_ for different shedding load distributions. We scanned across different parameter pairs (mean, standard deviation in days) for the SLD from time since infection. For the city of Zurich, the R_ww_ from N1 loads in wastewater was compared to that of confirmed cases in the catchment. For San Jose, we compared S gene concentrations to confirmed cases in Santa Clara County. The contour lines show SLD parameter pairs with equal coverage, in steps of 10% of the optimum value. In each comparison (grid-point) the 95% confidence intervals of R_cc_ and R_ww_ are based on 50 bootstrap replicates.

**Fig. S8:**
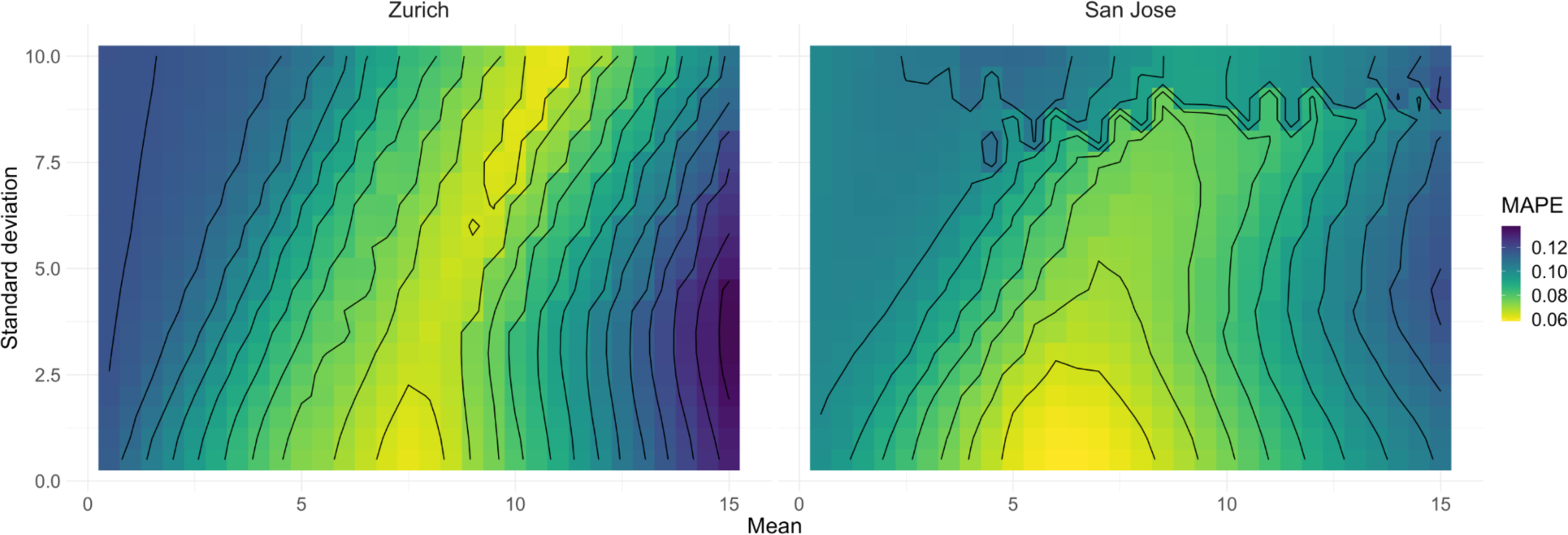
MAPE between R_cc_ and R_ww_ for different shedding load distributions. We scanned across different parameter pairs (mean, standard deviation in days) for the SLD from time since infection. For the city of Zurich, the R_ww_ from N1 loads in wastewater was compared to that of confirmed cases in the catchment. For San Jose, we compared S gene concentrations to confirmed cases in Santa Clara County. The contour lines show SLD parameter pairs with equal MAPE, in steps of 10% of the optimum value.

**Fig. S9:**
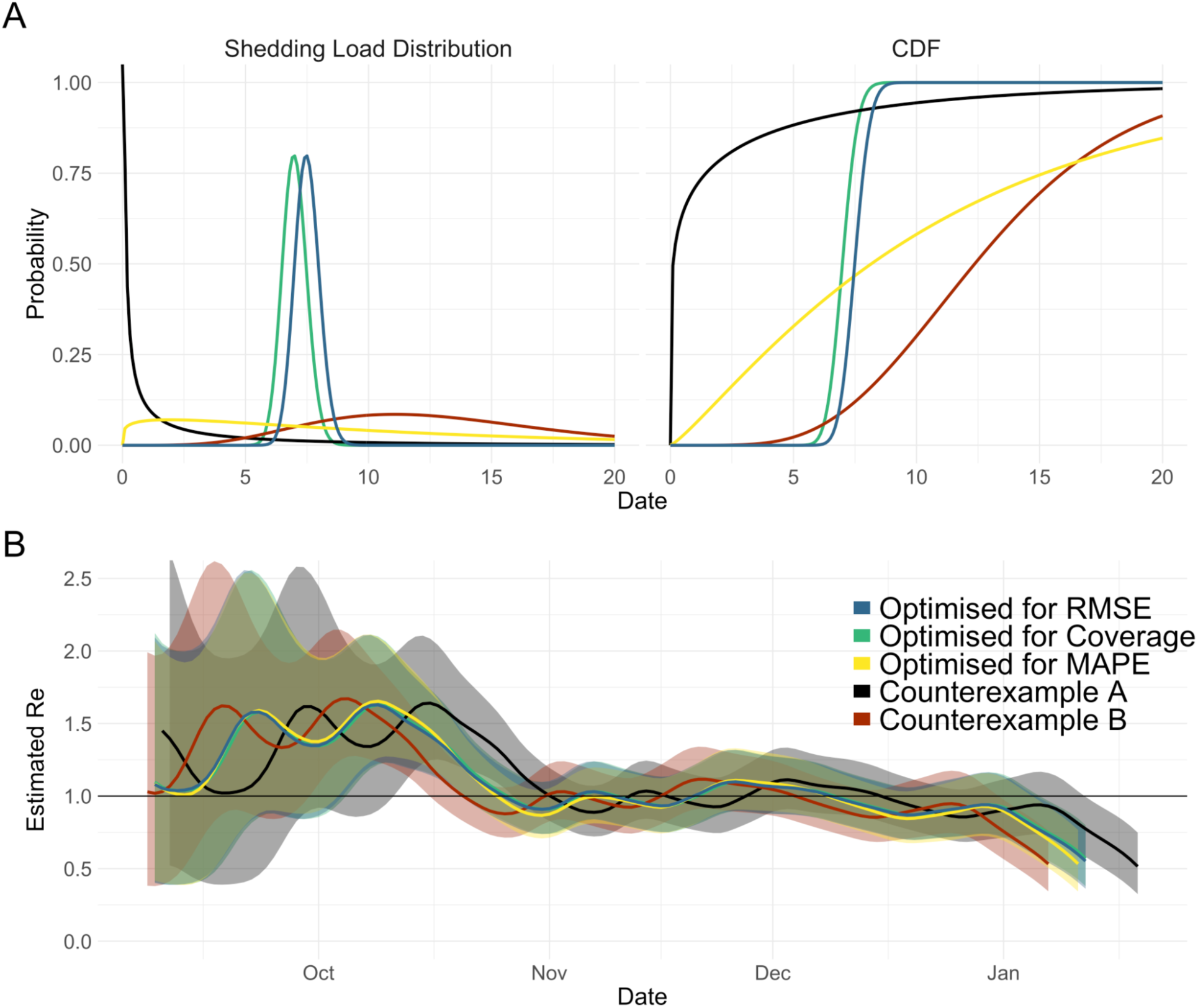
The optimal shedding load distributions for Zurich and two counterexamples. Counterexample A has a mean of 2 days and standard deviation of 5 days; Counterexample B a mean of 13 days, and a standard deviation of 5 days. **(A)** The SLDs and their associated cumulative distribution functions. Although the SLDs differ when optimized for RMSE, Coverage, or MAPE, the median (i.e. the time after which they reach half their overall probability mass) is very comparable. **(B)** The resulting R_ww_ estimates for Zurich are highly similar for RMSE, Coverage, and MAPE based optimal SLDs; but differ substantially for the two counterexamples. The illustrated 95% confidence intervals of R_ww_ are based on 50 bootstrap replicates.

**Fig. S10:**
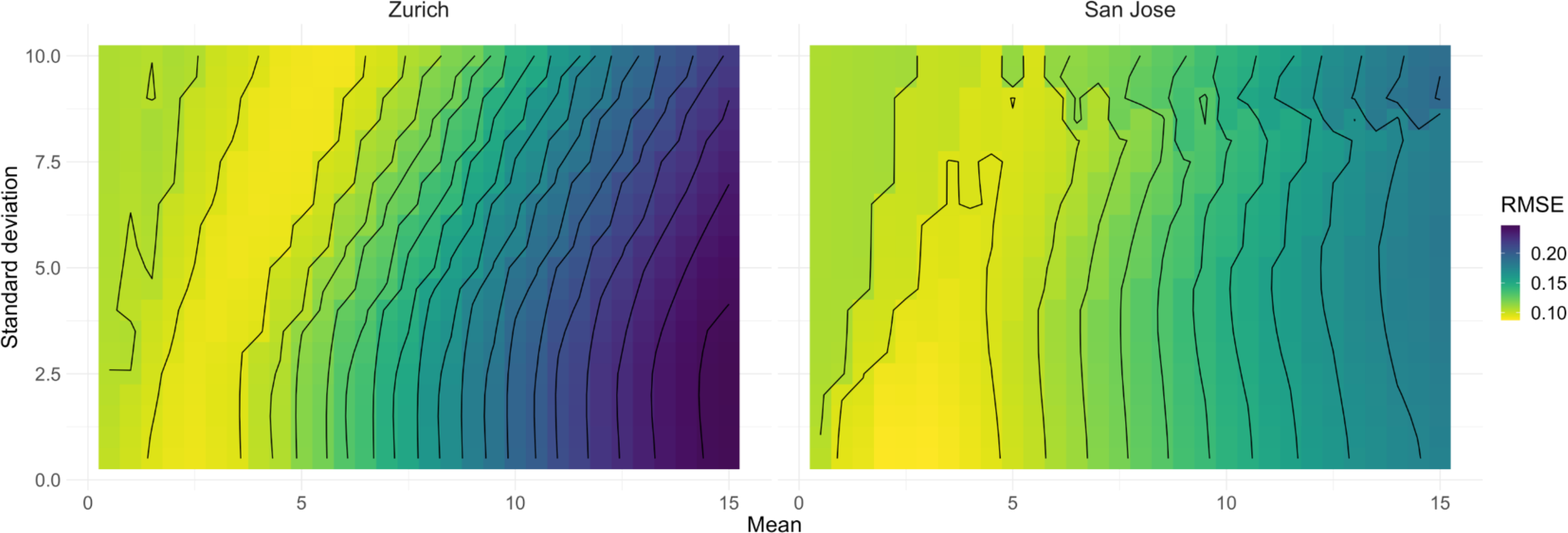
RMSE between R_cc_ and R_ww_ for different shedding load distributions from symptom onset. We scanned across different parameter pairs (mean, standard deviation in days) for the SLD from time since symptom onset. For the city of Zurich, the Re from N1 loads in wastewater was compared to that of confirmed cases in the catchment. For San Jose, we compared S gene concentrations to confirmed cases in Santa Clara County. The incubation period is assumed gamma distributed with mean 5.3, and standard deviation 3.2. The contour lines show SLD parameter pairs with equal RMSE, in steps of 10% of the optimum value.

**Fig. S11:**
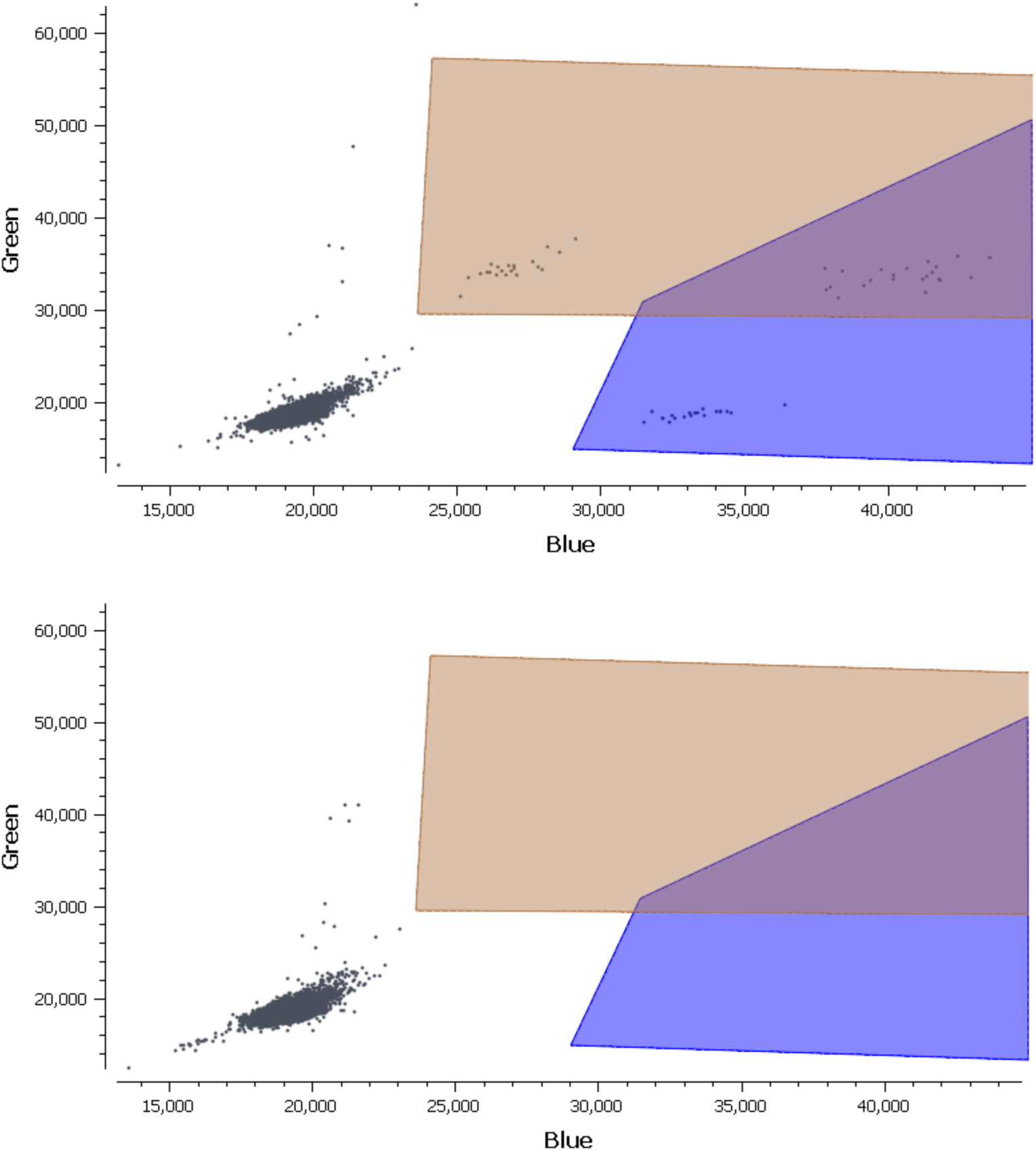
Fluorescence plots from the Stilla Naica Crystal Droplet PCR used for the wastewater. samples from Zurich collected after 23 September 2020 from positive (top) and negative (bottom) experimental results. Droplets positive for N1 (blue) and N2 (brown) and both N1 and N2 (purple). RPP30 markers, detectable as elevated fluorescence in Green channel, are included in the commercial assay used, but are not further analyzed.

**Fig. S12:**
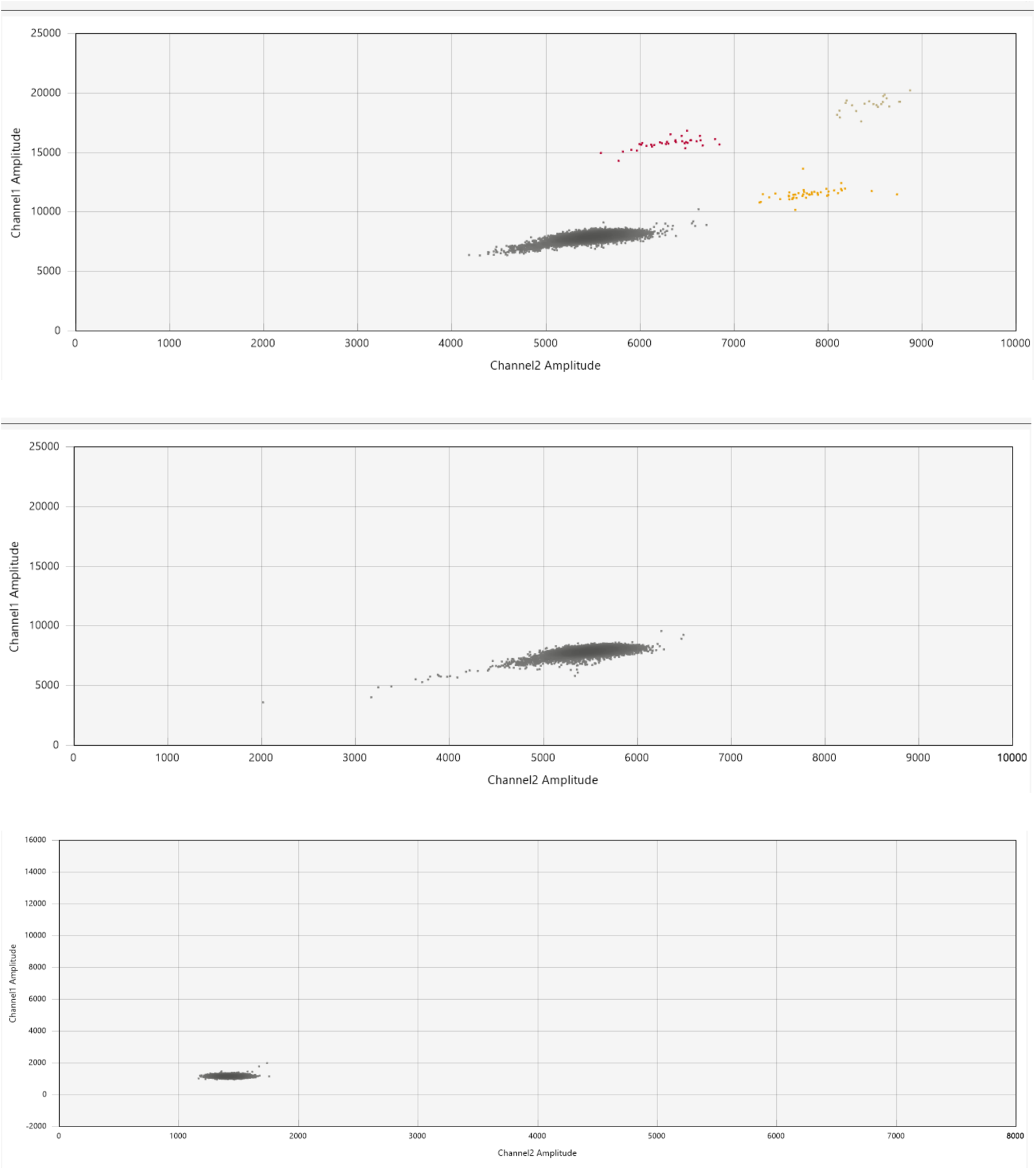
Fluorescence plots from the Bio-Rad QX200 used for the wastewater samples. Samples from Zurich collected from 03-20 September 2020 are shown in positive (top) and negative (middle) experimental results. Droplets positive for N1 (red) and N2 (yellow) and both N1 and N2 (brown) markers. Negative (bottom) experimental results are also shown for San Jose Corresponding positive experimental results are provided in the reference of the associated protocol ^30^.

**Table S1:**
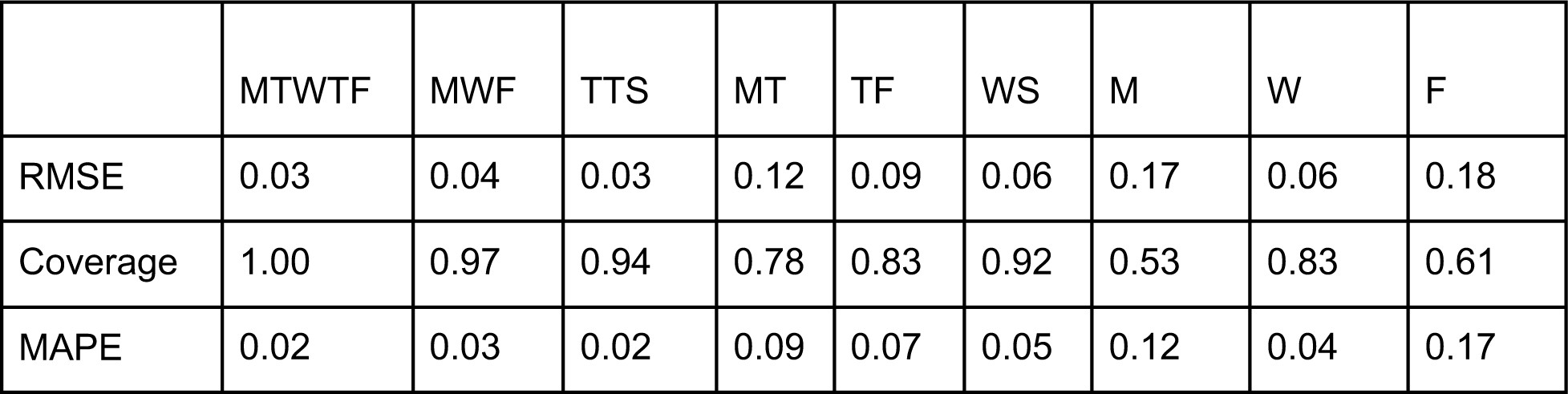
T**h**e **match between the daily and further subsampled R_ww_ traces for Zurich.** The data stems from measurements of the N1 marker for Zurich between 2020-11-22 and 2021-01-11. All traces are compared to the daily sampling; MTWTF corresponds to sampling during the working week, MWF Mon-Wed-Fri, TTS Tue-Thu-Sat, MT Mon-Thu, TF Tue-Fri, WS Wed-Sat, M Monday, W Wednesday, F Friday.

|  | MTWTF | MWF | TTS | MT | TF | WS | M | W | F |
| --- | --- | --- | --- | --- | --- | --- | --- | --- | --- |
| RMSE | 0.03 | 0.04 | 0.03 | 0.12 | 0.09 | 0.06 | 0.17 | 0.06 | 0.18 |
| Coverage | 1.00 | 0.97 | 0.94 | 0.78 | 0.83 | 0.92 | 0.53 | 0.83 | 0.61 |
| MAPE | 0.02 | 0.03 | 0.02 | 0.09 | 0.07 | 0.05 | 0.12 | 0.04 | 0.17 |

**Table S2:** T**h**e **match between the daily and further subsampled R_ww_ traces for San Jose.** The data stems from the S gene measurements for San Jose. All traces are compared to the daily sampling; MTWTF corresponds to sampling during the working week, MWF Mon-Wed-Fri, TTS Tue-Thu-Sat, MT Mon-Thu, TF Tue-Fri, WS Wed-Sat, M Monday, W Wednesday, F Friday.

|  | MTWTF | MWF | TTS | MT | TF | WS | M | W | F |
| --- | --- | --- | --- | --- | --- | --- | --- | --- | --- |
| RMSE | 0.05 | 0.09 | 0.08 | 0.15 | 0.06 | 0.16 | 0.13 | 0.13 | 0.08 |
| Coverage | 0.95 | 0.88 | 0.91 | 0.83 | 0.93 | 0.60 | 0.76 | 0.68 | 0.84 |
| MAPE | 0.05 | 0.08 | 0.07 | 0.15 | 0.06 | 0.14 | 0.11 | 0.11 | 0.07 |

**Table S3:** P**a**rameters **of the optimal shedding distribution from symptom onset.** We scanned across different (mean, standard deviation) parameter pairs for the SLD from time since symptom onset. For the city of Zurich, the R_ww_ from N1 loads in wastewater was compared to that of confirmed cases in the catchment. For San Jose, we compared S gene concentrations to confirmed cases in Santa Clara County. For all RMSE values of the scan see Fig. S9. All parameters are in units of days. For the coverage, the 95% confidence intervals of R_cc_ and R_ww_ were based on 50 bootstrap replicates in each comparison.

| Comparison Method | Optimal pair (mean; sd) | Mean within 10% of the optimum | SD within 10% of the optimum |
| --- | --- | --- | --- |
| RMSE (Zurich) | (5.5; 10) | [1.5, 7] | [0.5, 10] |
| Coverage (Zurich) | (3.5; 5) | [1.5, 8] | [0.5, 10] |
| MAPE (Zurich) | (5.5; 10) | [2, 6] | [0.5, 10] |
| RMSE (San Jose) | (3; 0.5) | [1, 5] | [0.5, 9] |
| Coverage (San Jose) | (0.5; 0.5) | [0.5, 6] | [0.5, 10] |
| MAPE (San Jose) | (1; 0.5) | [0.5, 3] | [0.5, 10] |

**Table S4:**
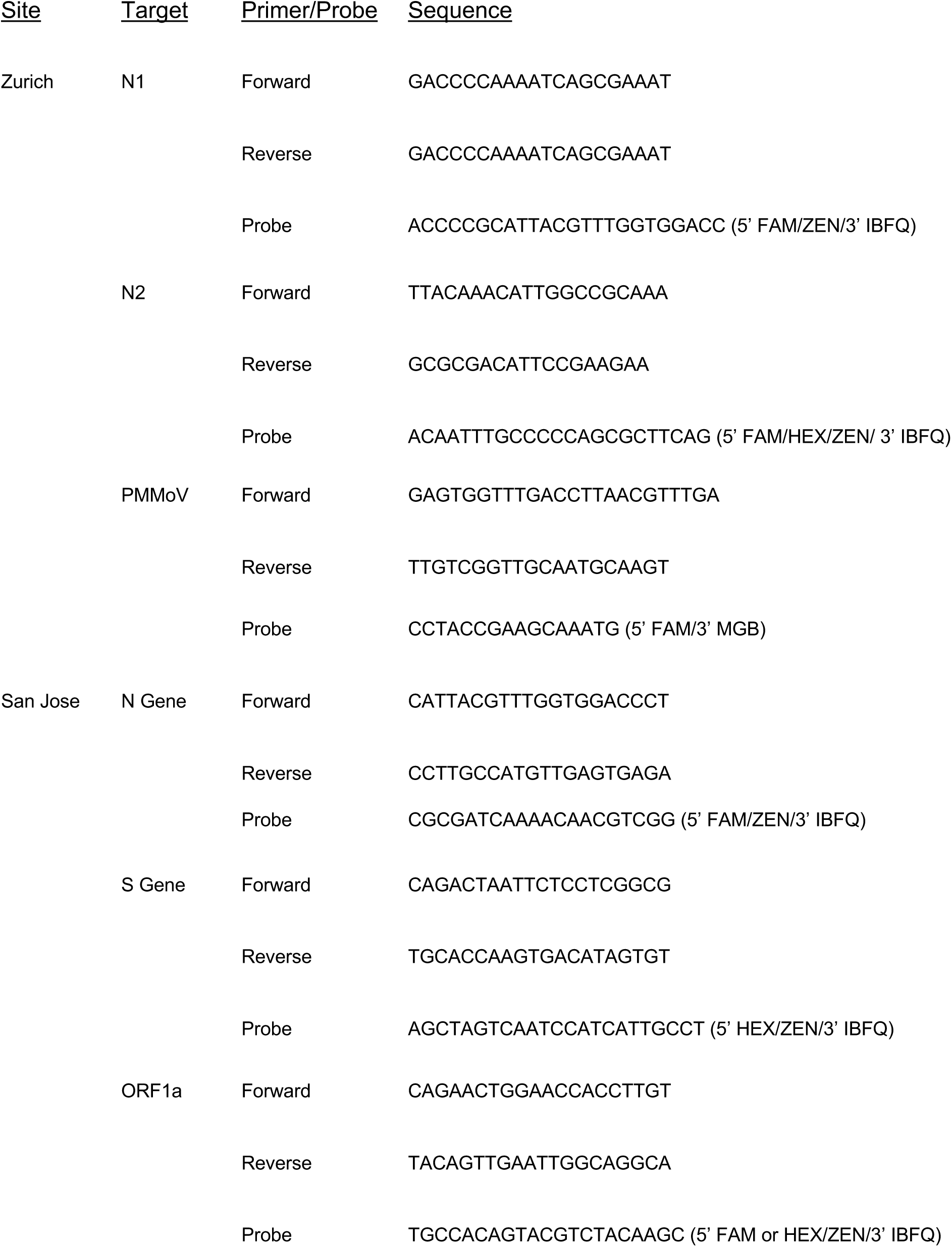
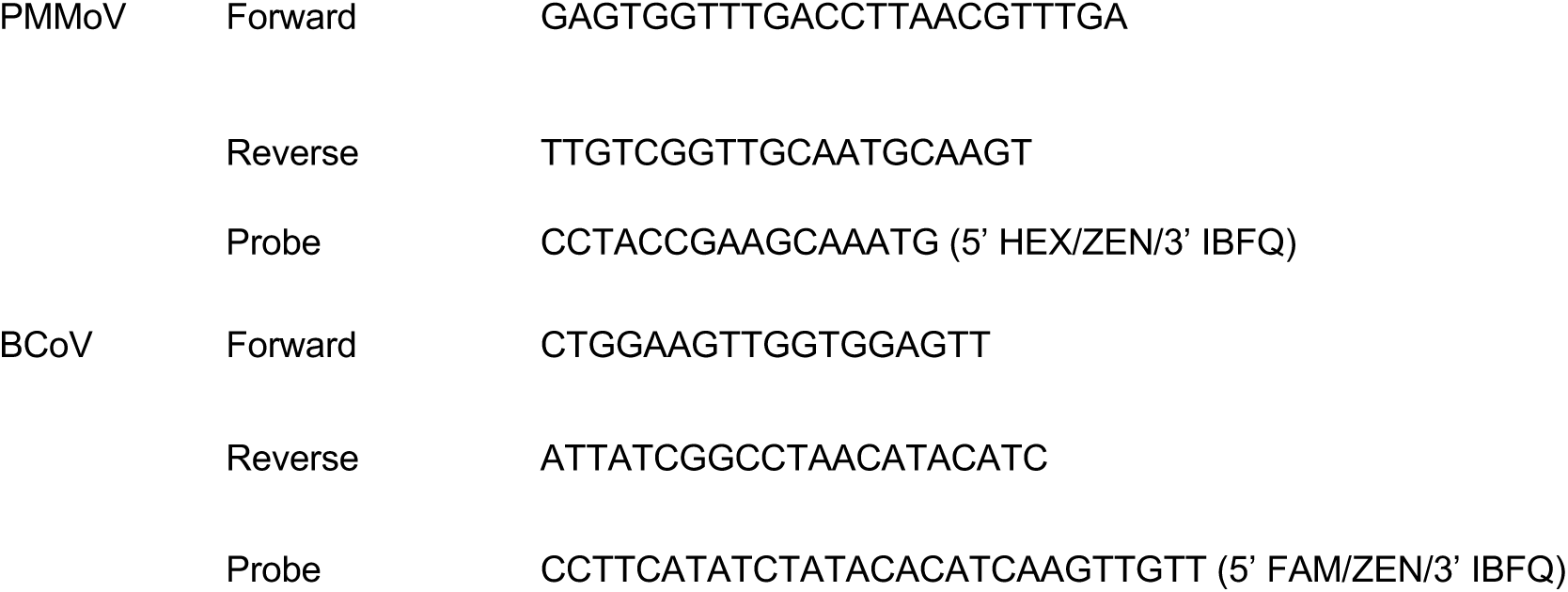
Primer and Probes

**Table S5:**
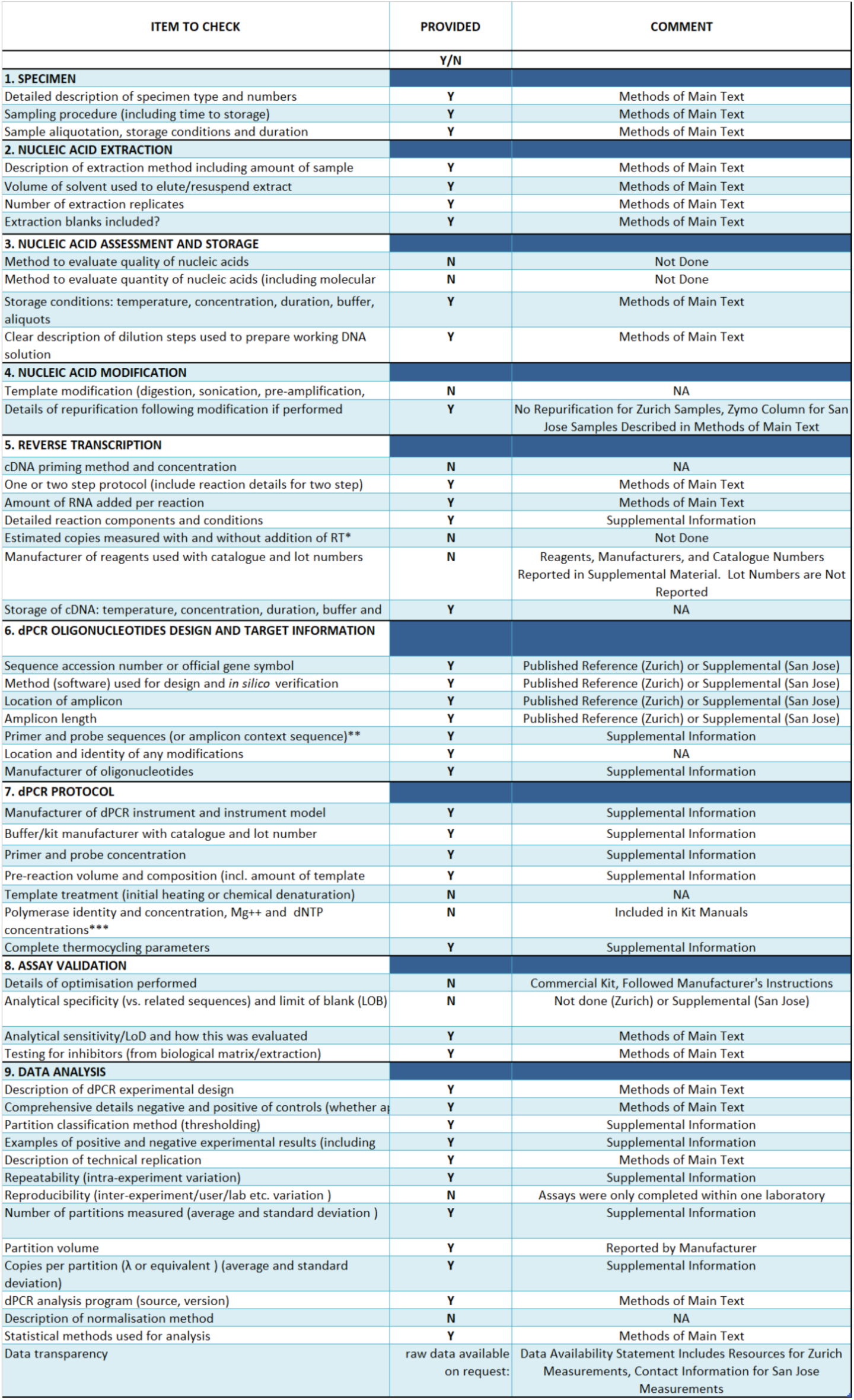
dMIQE Checklist ^63^ for RT-dPCR assays targeting SARS-CoV-2 gene markers N1 and N2 (Zurich Site) and S, N, and Orf1a (San Jose Site). Checklist is not included for other RT-qPCR (PMMoV, Zurich Site) or RT-dPCR (BCoV, PMMoV, San Jose) Assays.

## Supplemental Results

### Quality Control

#### Zurich, Switzerland

One sample (11 October) was removed from analysis because the dilute sample (1:10) was below LOQ and the undiluted sample was inhibited, as defined by recovery of less than 80% of the synthetic SARS-CoV-2 RNA added in.

PMMoV concentrations were obtained for all dates except October 4. Mean (standard deviation) PMMoV loads were 16.5 (0.12) log10 (gc/day). All PMMoV loads fell within 3 standard deviations of the mean, consistent with a normal distribution, except on 29 October (16.1 log10 gc/day). The sample was subsequently removed from further analysis.

#### San Jose, California

*USA.* From San Jose Wastewater Treatment Plant (San Jose, California, USA), daily samples were collected and processed throughout. PMMoV concentrations were mean (standard deviation) 8.9 (0.20) log10 (gc/g-dry weight). On two days (03 January 2021, 18 February 2021), PMMoV concentrations exceeded the mean plus three times the standard deviation. On one day (19 March 2021), PMMoV concentrations fell below the mean minus three times the standard deviation. These three samples were excluded from further analysis. All samples met criteria for inclusion based on BCoV concentrations, which were all greater than 10% of the expected concentrations based on the amount added.

## Supplemental Methods

### Managing PCR inhibition in San Jose sludge samples

We diluted the solids in DNA/RNA Shield (Zymo Research, Irvine, CA, USA) (hereafter referred to as “diluent”) prior to RNA extraction, inhibitor removal, and RT-dPCR, as described in the main text. This dilution was necessary as direct RNA extraction from the solids contained inhibitors so dilution of the RNA template prior to RT-dPCR was required ^15^.To determine an optimal concentration of solids to add to the diluent (mg wet weight / mL diluent) to reduce inhibition, but retain reasonable sensitivity, we quantified SARS-CoV-2 RNA in solutions with solids concentrations from 7.5 mg/mL to 150 mg/mL, where we used the exact methods described in the main text. We aimed to identify the highest concentrations of solids in solution that did not show inhibition. Representative results from two different samples from San Jose are shown in Figure S13 where concentrations of SARS-CoV-2 RNA per g dry weight of solids is shown for different starting concentrations of solids in diluent. Note that the numbers on the y-axis are directly comparable between solutions, as we account for the different mass of solids in the dimensional analysis to derive the SARS-CoV-2 RNA concentration. The relatively lower concentrations for SARS-CoV-2 RNA for solutions of 150 mg/mL suggest the presence of inhibition at this relatively high solids concentration. Concentrations are relatively similar among the lower concentration solutions suggesting that starting at 75 mg/mL, inhibition is mostly alleviated. Only one gene target from one of the samples (the N gene in sample A shown in the top graph) showed slightly higher concentrations in more dilute solutions. Given these results we therefore moved forward using 75 mg/mL for the method.

**Figure S13.**
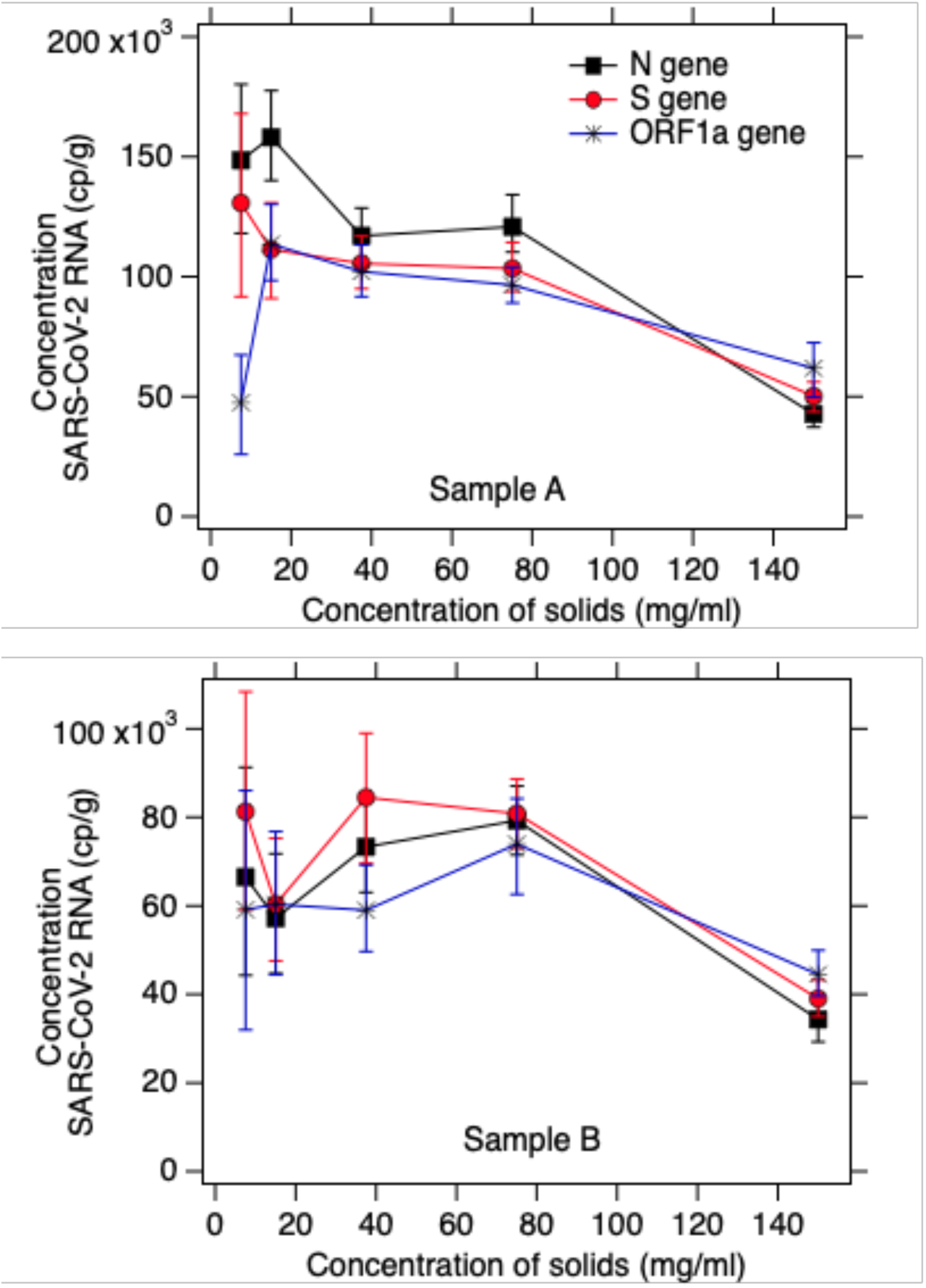
Representative results from inhibition titration experiments with two samples from San Jose. The x-axis shows the concentration of solids (wet weight) added to the DNA/RNA Shield diluent in units of mg/mL. The y-axis shows the concentration of the three SARS-CoV-2 RNA targets in the solids in units of gc/g dry weight. Error bars represent standard deviations as total error from the dPCR instrument’s software.

## Notes

### Competing Interest Statement

The authors have declared no competing interest.

### Author Declarations

No ethics approval was necessary

## References

1. Gostic KM, McGough L, Baskerville EB, et al. Practical considerations for measuring the effective reproductive number, R t. PLoS Comput Biol. 2020;16(12):e1008409.

2. Huisman JS, Scire J, Angst DC, Neher RA, Bonhoeffer S, Stadler T. Estimation and worldwide monitoring of the effective reproductive number of SARS-CoV-2. medrxiv. Published online 2020. https://www.medrxiv.org/content/10.1101/2020.11.26.20239368v1.abstract

3. Wallinga J, Teunis P. Different epidemic curves for severe acute respiratory syndrome reveal similar impacts of control measures. Am J Epidemiol. 2004;160(6):509–516.

4. Cori A, Ferguson NM, Fraser C, Cauchemez S. A new framework and software to estimate time-varying reproduction numbers during epidemics. Am J Epidemiol. 2013;178(9):1505–1512.

5. Brauner JM, Mindermann S, Sharma M, et al. Inferring the effectiveness of government interventions against COVID-19. Science. 2021;371(6531). doi:10.1126/science.abd9338

6. Der Schweizerische Bundesrat. Verordnung über Massnahmen in Der Besonderen Lage Zur Bekämpfung Der Covid-19-Epidemie.; 2020.

7. Anderson R, Donnelly C, Hollingsworth D, et al. Reproduction Number (R) and Growth Rate (r) of the COVID-19 Epidemic in the UK: Methods of Estimation, Data Sources, Causes of Heterogeneity, and Use as a Guide in Policy Formulation. The Royal Society; 2020.

8. Yabe T, Tsubouchi K, Fujiwara N, Wada T, Sekimoto Y, Ukkusuri SV. Non-compulsory measures sufficiently reduced human mobility in Tokyo during the COVID-19 epidemic. Sci Rep. 2020;10(1):18053.

9. Pan A, Liu L, Wang C, et al. Association of Public Health Interventions With the Epidemiology of the COVID-19 Outbreak in Wuhan, China. JAMA. 2020;323(19):1915–1923.

10. Flaxman S, Mishra S, Gandy A, et al. Estimating the effects of non-pharmaceutical interventions on COVID-19 in Europe. Nature. 2020;584(7820):257-261.

11. Soltesz K, Gustafsson F, Timpka T, et al. The effect of interventions on COVID-19. Nature. 2020;588(7839):E26-E28.

12. Rossen LM, Branum AM, Ahmad FB, Sutton P, Anderson RN. Excess Deaths Associated with COVID-19, by Age and Race and Ethnicity — United States, January 26–October 3, 2020. MMWR Morbidity and Mortality Weekly Report. 2020;69(42):1522–1527. doi:10.15585/mmwr.mm6942e2

13. Peccia J, Zulli A, Brackney DE, et al. Measurement of SARS-CoV-2 RNA in wastewater tracks community infection dynamics. Nat Biotechnol. 2020;38(10):1164–1167.

14. Karthikeyan S, Ronquillo N, Belda-Ferre P, et al. High-Throughput Wastewater SARS-CoV-2 Detection Enables Forecasting of Community Infection Dynamics in San Diego County. mSystems. 2021;6(2). doi:10.1128/mSystems.00045-21

15. Graham KE, Loeb SK, Wolfe MK, et al. SARS-CoV-2 RNA in Wastewater Settled Solids Is Associated with COVID-19 Cases in a Large Urban Sewershed. Environ Sci Technol. 2021;55(1):488–498.

16. Medema G, Heijnen L, Elsinga G, Italiaander R, Brouwer A. Presence of SARS-Coronavirus-2 RNA in Sewage and Correlation with Reported COVID-19 Prevalence in the Early Stage of the Epidemic in The Netherlands. Environ Sci Technol Lett. 2020;7(7):511–516.

17. Agrawal S, Orschler L, Lackner S. Long-term monitoring of SARS-CoV-2 RNA in wastewater of the Frankfurt metropolitan area in Southern Germany. Sci Rep. 2021;11(1):5372.

18. Arora S, Nag A, Sethi J, et al. Sewage surveillance for the presence of SARS-CoV-2 genome as a useful wastewater based epidemiology (WBE) tracking tool in India. Water Sci Technol. 2020;82(12):2823–2836.

19. Haramoto E, Malla B, Thakali O, Kitajima M. First environmental surveillance for the presence of SARS-CoV-2 RNA in wastewater and river water in Japan. Sci Total Environ. 2020;737:140405.

20. Kaplan EH, Wang D, Wang M, Malik AA, Zulli A, Peccia J. Aligning SARS-CoV-2 indicators via an epidemic model: application to hospital admissions and RNA detection in sewage sludge. Health Care Manag Sci. Published online October 28, 2020. doi:10.1007/s10729-020-09525-1

21. McMahan CS, Self S, Rennert L, et al. COVID-19 Wastewater Epidemiology: A Model to Estimate Infected Populations. MedRxiv. Published online 2020. https://www.medrxiv.org/content/10.1101/2020.11.05.20226738v1.abstract

22. Fernandez-Cassi X, Scheidegger A, Bänziger C, et al. Wastewater monitoring outperforms case numbers as a tool to track COVID-19 incidence dynamics when test positivity rates are high. doi:10.1101/2021.03.25.21254344

23. Wölfel R, Corman VM, Guggemos W, et al. Virological assessment of hospitalized patients with COVID-2019. Nature. 2020;581(7809):465-469.

24. Han MS, Seong MW, Kim N, et al. Viral RNA Load in Mildly Symptomatic and Asymptomatic Children with COVID-19, Seoul, South Korea. Emerg Infect Dis. 2020;26(10):2497–2499.

25. Benefield AE, Skrip LA, Clement A, Althouse RA, Chang S, Althouse BM. SARS-CoV-2 viral load peaks prior to symptom onset: a systematic review and individual-pooled analysis of coronavirus viral load from 66 studies. medRxiv. Published online 2020. https://www.medrxiv.org/content/10.1101/2020.09.28.20202028v1.abstract

26. Li Q, Guan X, Wu P, et al. Early Transmission Dynamics in Wuhan, China, of Novel Coronavirus–Infected Pneumonia. N Engl J Med. 2020;382(13):1199–1207.

27. Symonds EM, Nguyen KH, Harwood VJ, Breitbart M. Pepper mild mottle virus: A plant pathogen with a greater purpose in (waste)water treatment development and public health management. Water Res. 2018;144:1–12.

28. Topol A, Wolfe M, White B, Wigginton K, Boehm A. High Throughput pre-analytical processing of wastewater settled solids for SARS-CoV-2 RNA analyses v1 (protocols.io.btyqnpvw). *protocols.io*. Published online 2021. doi:10.17504/protocols.io.btyqnpvw

29. Topol A, Wolfe M, Wigginton K, White B, Boehm A. High Throughput RNA Extraction and PCR Inhibitor Removal of Settled Solids for Wastewater Surveillance of SARS-CoV-2 RNA v1 (protocols.io.btyrnpv6). protocols.io. Published online 2021. doi:10.17504/protocols.io.btyrnpv6

30. Topol A, Wolfe M, White B, Wigginton K, Boehm A. High Throughput SARS-COV-2, PMMOV, and BCoV quantification in settled solids using digital RT-PCR v1 (protocols.io.btywnpxe). protocols.io. Published online 2021. doi:10.17504/protocols.io.btywnpxe

31. Wolfe MK, Topol A, Knudson A, et al. High-Frequency, High-Throughput Quantification of SARS-CoV-2 RNA in Wastewater Settled Solids at Eight Publicly Owned Treatment Works in Northern California Shows Strong Association with COVID-19 Incidence. mSystems. 2021;6(5):e0082921.

32. Zhu K, Suttner B, Pickering A, Konstantinidis KT, Brown J. A novel droplet digital PCR human mtDNA assay for fecal source tracking. Water Res. 2020;183:116085.

33. Huggett JF, Novak T, Garson JA, et al. Differential susceptibility of PCR reactions to inhibitors: an important and unrecognised phenomenon. BMC Res Notes. 2008;1:70.

34. Haramoto E, Kitajima M, Kishida N, et al. Occurrence of pepper mild mottle virus in drinking water sources in Japan. Appl Environ Microbiol. 2013;79(23):7413–7418.

35. Zhang T, Breitbart M, Lee WH, et al. RNA Viral Community in Human Feces: Prevalence of Plant Pathogenic Viruses. PLoS Biol. 2005;4(1):e3.

36. Linton NM, Kobayashi T, Yang Y, et al. Incubation period and other epidemiological characteristics of 2019 novel Coronavirus infections with right truncation: A statistical analysis of publicly available case data. J Clin Med Res. 2020;9(2):538.

37. Cori A, Cauchemez S, Ferguson NM, et al. EpiEstim: estimate time varying reproduction numbers from epidemic curves. R package version. Published online 2019:2–2.

38. Feng S, Roguet A, McClary-Gutierrez JS, et al. Evaluation of sampling frequency and normalization of SARS-CoV-2 wastewater concentrations for capturing COVID-19 burdens in the community. medRxiv. Published online 2021. https://www.medrxiv.org/content/10.1101/2021.02.17.21251867v2.abstract

39. Kitajima M, Ahmed W, Bibby K, et al. SARS-CoV-2 in wastewater: State of the knowledge and research needs. Sci Total Environ. 2020;739:139076.

40. Cevik M, Tate M, Lloyd O, Maraolo AE, Schafers J, Ho A. SARS-CoV-2, SARS-CoV, and MERS-CoV viral load dynamics, duration of viral shedding, and infectiousness: a systematic review and meta-analysis. Lancet Microbe. 2021;2(1):e13–e22.

41. Zheng S, Fan J, Yu F, et al. Viral load dynamics and disease severity in patients infected with SARS-CoV-2 in Zhejiang province, China, January-March 2020: retrospective cohort study. BMJ. 2020;369:m1443.

42. Walsh KA, Jordan K, Clyne B, et al. SARS-CoV-2 detection, viral load and infectivity over the course of an infection. J Infect. 2020;81(3):357–371.

43. Hoffmann T, Alsing J. Faecal shedding models for SARS-CoV-2 RNA amongst hospitalised patients and implications for wastewater-based epidemiology. medRxiv. Published online 2021. https://www.medrxiv.org/content/10.1101/2021.03.16.21253603v1.abstract

44. Liu Y, Yan LM, Wan L, et al. Viral dynamics in mild and severe cases of COVID-19. Lancet Infect Dis. 2020;20(6):656–657.

45. Zhou C, Zhang T, Ren H, et al. Impact of age on duration of viral RNA shedding in patients with COVID-19. Aging. 2020;12(22):22399–22404.

46. Parag KV. Improved estimation of time-varying reproduction numbers at low case incidence and between epidemic waves. *medRxiv*. Published online 2020. https://www.medrxiv.org/content/10.1101/2020.09.14.20194589v1.abstract

47. Abbott S, Hellewell J, Thompson RN, et al. Estimating the time-varying reproduction number of SARS-CoV-2 using national and subnational case counts. Wellcome Open Research. 2020;5:112. doi:10.12688/wellcomeopenres.16006.1

48. Stringhini S, Wisniak A, Piumatti G, et al. Seroprevalence of anti-SARS-CoV-2 IgG antibodies in Geneva, Switzerland (SEROCoV-POP): a population-based study. Lancet. 2020;396(10247):313–319.

49. Stringhini S, Zaballa ME, Perez-Saez J, et al. Seroprevalence of anti-SARS-CoV-2 antibodies after the second pandemic peak. The Lancet Infectious Diseases. Published online 2021. doi:10.1016/s1473-3099(21)00054-2

50. Heaton KW, Radvan J, Cripps H, Mountford RA, Braddon FE, Hughes AO. Defecation frequency and timing, and stool form in the general population: a prospective study. Gut. 1992;33(6):818–824.

51. Thomas KV, Amador A, Baz-Lomba JA, Reid M. Use of Mobile Device Data To Better Estimate Dynamic Population Size for Wastewater-Based Epidemiology. Environ Sci Technol. 2017;51(19):11363–11370.

52. Kantor RS, Nelson KL, Greenwald HD, Kennedy LC. Challenges in Measuring the Recovery of SARS-CoV-2 from Wastewater. Environ Sci Technol. 2021;55(6):3514–3519.

53. de Oliveira LC, Torres-Franco AF, Lopes BC, et al. Viability of SARS-CoV-2 in river water and wastewater at different temperatures and solids content. Water Res. 2021;195:117002.

54. Bivins A, Greaves J, Fischer R, et al. Persistence of SARS-CoV-2 in Water and Wastewater. Environ Sci Technol Lett. 2020;7(12):937–942.

55. Hokajärvi AM, Rytkönen A, Tiwari A, et al. The detection and stability of the SARS-CoV-2 RNA biomarkers in wastewater influent in Helsinki, Finland. Sci Total Environ. 2021;770:145274.

56. Gerrity D, Papp K, Stoker M, Sims A, Frehner W. Early-pandemic wastewater surveillance of SARS-CoV-2 in Southern Nevada: Methodology, occurrence, and incidence/prevalence considerations. Water Res X. 2021;10:100086.

57. Jahn K, Dreifuss D, Topolsky I, et al. Detection of SARS-CoV-2 variants in Switzerland by genomic analysis of wastewater samples. medRxiv. Published online 2021. https://www.medrxiv.org/content/10.1101/2021.01.08.21249379v1.abstract

58. Crits-Christoph A, Kantor RS, Olm MR, et al. Genome Sequencing of Sewage Detects Regionally Prevalent SARS-CoV-2 Variants. MBio. 2021;12(1). doi:10.1128/mBio.02703-20

59. Martin J, Klapsa D, Wilton T, et al. Tracking SARS-CoV-2 in Sewage: Evidence of Changes in Virus Variant Predominance during COVID-19 Pandemic. Viruses. 2020;12(10). doi:10.3390/v12101144

60. Brinkman NE, Fout GS, Keely SP. Retrospective Surveillance of Wastewater To Examine Seasonal Dynamics of Enterovirus Infections. mSphere. 2017;2(3). doi:10.1128/mSphere.00099-17

61. Kazama S, Masago Y, Tohma K, et al. Temporal dynamics of norovirus determined through monitoring of municipal wastewater by pyrosequencing and virological surveillance of gastroenteritis cases. Water Res. 2016;92:244–253.

62. McCall C, Wu H, Miyani B, Xagoraraki I. Identification of multiple potential viral diseases in a large urban center using wastewater surveillance. Water Res. 2020;184:116160.

63. dMIQE Group, Huggett JF. The Digital MIQE Guidelines Update: Minimum Information for Publication of Quantitative Digital PCR Experiments for 2020. Clin Chem. 2020;66(8):1012–1029.

